# Advancing Precision Health Discovery in a Genetically Diverse Health System

**DOI:** 10.1101/2025.06.11.25329386

**Authors:** Roni Haas, Michael P. Margolis, Angela Wei, Takafumi N Yamaguchi, Jeffrey Feng, Thai Tran, Veronica Tozzo, Katelyn J. Queen, Mohammed Faizal Eeman Mootor, Vishakha Patil, Michael E. Broudy, Paul Tung, Shafiul Alam, Danielle B. Martinez, Yash Patel, Christa Caggiano, Nicole Zeltser, Rupert Hugh-White, Jaron Arbet, Ruhollah Shemirani, Mao Tian, Prapti Thapaliya, Lora Eloyan, Lawrence O. Chen, Sandra Lapinska, Maryam Ariannejad, Clara Lajonchere, UCLA Precision Health Data Discovery Repository Working Group, UCLA Precision Health ATLAS Working Group, UCLA Health IT HPC Team, Regeneron Genetics Center, Eimear E. Kenny, Bogdan Pasaniuc, Alex A. T. Bui, Valerie A. Arboleda, Timothy S. Chang, Noah Zaitlen, Paul T. Spellman, Paul C. Boutros, Daniel H. Geschwind

**Author notes:** ^18^ These authors contributed equally (Roni Haas, Michael P. Margolis, Angela Wei). ^19^ Senior authors (Paul T. Spellman, Paul C. Boutros, Daniel H. Geschwind). ^20^ Lead contact (Daniel H. Geschwind). **Correspondence:** (R.H.), (P.T.S.), (P.C.B.), (D.H.G.).

## Abstract

Linking genetic data with electronic health records in hospital biobanks promises to advance precision medicine, but limited ancestral diversity constrains discovery and generalizability. We analyzed 93,936 participants from the UCLA ATLAS Community Health Initiative to inform disease prevalence and genetic risk across five continental and 36 fine-scale ancestry groups. We discovered numerous unreported gene-phenotype associations, including *FN3K* with intestinal disaccharidase deficiency in Europeans and admixed Americans. Polygenic scores (PGS) robustly predicted common diseases, with effects markedly diminished in non-Europeans. Furthermore, we reduced the pronounced European bias in curated clinical variants using computational predictors, uncovering unreported disease-gene associations, including *ANKZF1* and peripheral vascular disease in AFR. Longitudinal data revealed that semaglutide efficacy varies across ancestries, is associated with PGS for type 2 diabetes, and is modulated by genetic variation in *PTPRU*. These findings illustrate how ancestrally diverse biobanks from a single health system yield robust disease associations and pharmacogenomic insights.

## Introduction

The integration of electronic health records (EHRs) with genetic data is transforming biomedical research, offering unprecedented opportunities for preventing and managing common medical conditions^1^. Longitudinal sampling, linked with genetic and environmental data, provides advantages over standard cohort-driven research^2^. This has fueled the creation of nationwide biobanks, including the UK Biobank (UKBB) ^3,4^, All of Us (AoU)^5^, FinnGen^6^ and Taiwan Biobank^7^, as well as several large-scale academic biobanks, such as Mt. Sinai’s BioMe^8^, Vanderbilt’s BioVU^9^, Geissinger’s MyCode^10^, and the Michigan Genomics Initiative^11^. Integration of these biobanks has permitted innovative collaborative efforts, such as the eMERGE consortium^12^, COVID-19 host genomics initiative^13^, and the Global Biobank Initiative^1^.

Although these efforts have substantially advanced genetic and biomedical discovery, their concentration on participants of European (EUR) ancestry limits generalizability^14–18^. As PGS are validated for clinical use, the importance of measuring their accuracy in diverse populations grows; recent analyses show a continuous relationship between ancestral distance from the reference population and the utility of PGS^17^. Similarly, rare genetic variation has substantial ancestry-specific effects and distribution; for example, in African Americans, *APOE4* alleles have reduced impact on Alzheimer’s disease risk^19^ and rare protective *PCSK9* variants are more prevalent^20^. The interpretation of clinically-relevant rare variation is further hampered by a bias towards EUR variants in genetic databases^21,22^. Including non-EUR populations reveals substantial disparities in clinically-relevant rare variant frequencies^23^ and increases statistical power for discovery^24,25^. Thus, greater ancestral diversity in biobanks with detailed medical records strengthens efforts to advance precision health^14,26,27^.

Here, we analyzed data from the UCLA ATLAS Community Health Initiative, linking EHR with genomic information for 92,164 participants with array genotyping and 61,797 with whole-exome sequencing (WES). ATLAS reflects Los Angeles’ ancestral diversity within a single health system^28–30^, which reduces confounding from differing clinical practices and enables robust cross-population comparisons. We performed phenome-wide associations using common and rare variants within continental (broad-scale) and sub-continental (fine-scale) ancestral groups, and quantified disease diagnoses and genetic risk across these groups. Using longitudinal EHR data, we use semaglutide as a case study to identify genetic factors that alter drug efficacy. We identified multiple unreported ancestry-specific risk associations, further demonstrating the utility of ancestral diversity for more equitable, personalized medicine research (**Figure 1a**, schematic overview).

**Figure 1.**
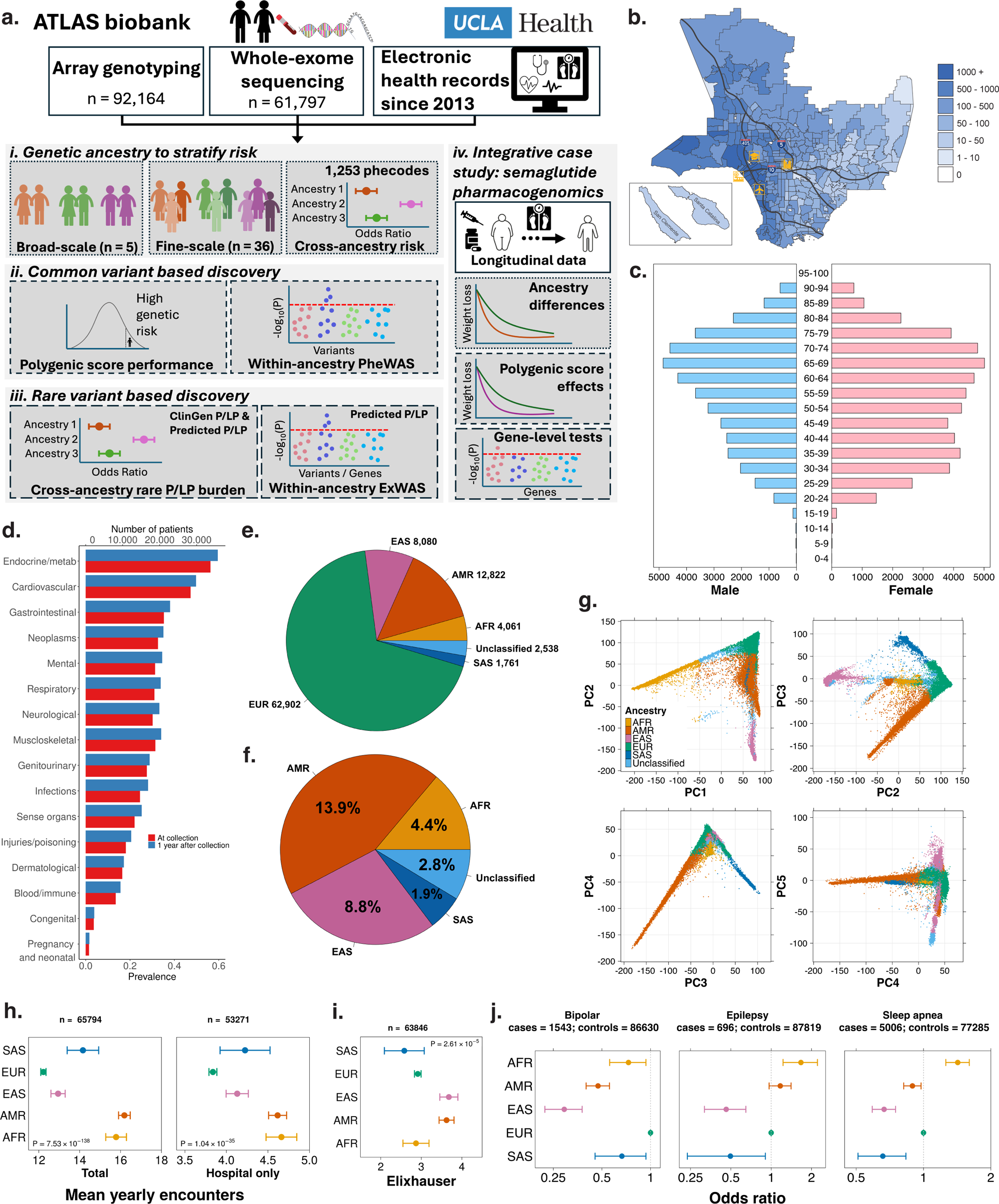
Overview of the UCLA ATLAS biobank. **a.** Schematic workflow**. b.** Choropleth map of ATLAS participants within Los Angeles County, with major highways and landmarks labelled. **c.** Distribution of ATLAS participants by age and sex. **d.** Prevalence of phecode groups in the UCLA ATLAS population at the time of collection and within one year of the ATLAS launch date. **e.** Genetic ancestry sample sizes. **f**. Genetic ancestry fractions of non-European (EUR) populations. Percentages were calculated using all ATLAS populations, including EUR. **g.** Genetic principal components of ATLAS individuals. **h.** Mean yearly encounters vary across genetic ancestries. **i.** Comorbidity index varies across genetic ancestries. In **h-I**, ANCOVA was used to obtain P-values; the adjusted means and 95% confidence intervals are presented. **j.** Associations between clinical phenotypes and broad-scale ancestries using logistic regression.

## Results

### The demographic and clinical landscape in ATLAS

UCLA Health serves Los Angeles County, one of the world’s most ancestrally diverse metropolitan areas, with a population of 9.6 million. To date, the UCLA ATLAS initiative has consented ∼220,000 UCLA Health patients and has collected biomaterials from ∼130,000. ATLAS enrollment largely reflects the composition of UCLA Health patients, who are concentrated across Los Angeles (**Figure 1b**). The EHR, initiated in 2013, enables continuous longitudinal stratification of participants by disease states, with a mean and median of 8.6 and

7.9 years of participation per individual. As of November 2024, ATLAS genomic data included WES data from 61,797 participants and custom array genotyping using the Illumina Global Screening Array from 92,164 participants. Extensive quality control (QC) indicated high data quality (**Figure S1**). We leveraged these data to interrogate social and genetic factors that affect disease risk and health outcomes (**Figure 1a**).

Genotyped cohort demographics are summarized in **Table 1** and the **Supplementary Note**. Biobank participants were older in age and had a higher comorbidity index than non-biobank patients (3.3 *vs.* 1.7 mean Elixhauser index; 1-year post-collection), consistent with a higher number of clinical visits, which favors enrollment (controlled for data-completeness**, Methods**; **Supplementary Note**; **Figure 1c).** EHR-based phenotypes, including vital signs, disease diagnoses, and lab tests, were defined through harmonization procedures and subsequently validated (**Methods**; **Figure S2a-l**). Common disease domains were led by endocrine/metabolic, cardiovascular, gastrointestinal disorders and neoplasms, reflecting known global health challenges (**Figure 1d**; **Figure S2m-p**; **Supplementary Note**)^31–33^. The ATLAS EHR data includes a total of 71,739,582 lab tests (considering complete blood count [CBC], lipid, metabolic, hemoglobin A1c [HbA1c], and 25-hydroxyvitamin D panels), with a mean of 540 lab test results per participant (**Table S1**). Detailed prescription data show a total of 5,952,958 prescriptions; the 50 most frequently prescribed medications are listed in **Table S1**.

**Table 1.**
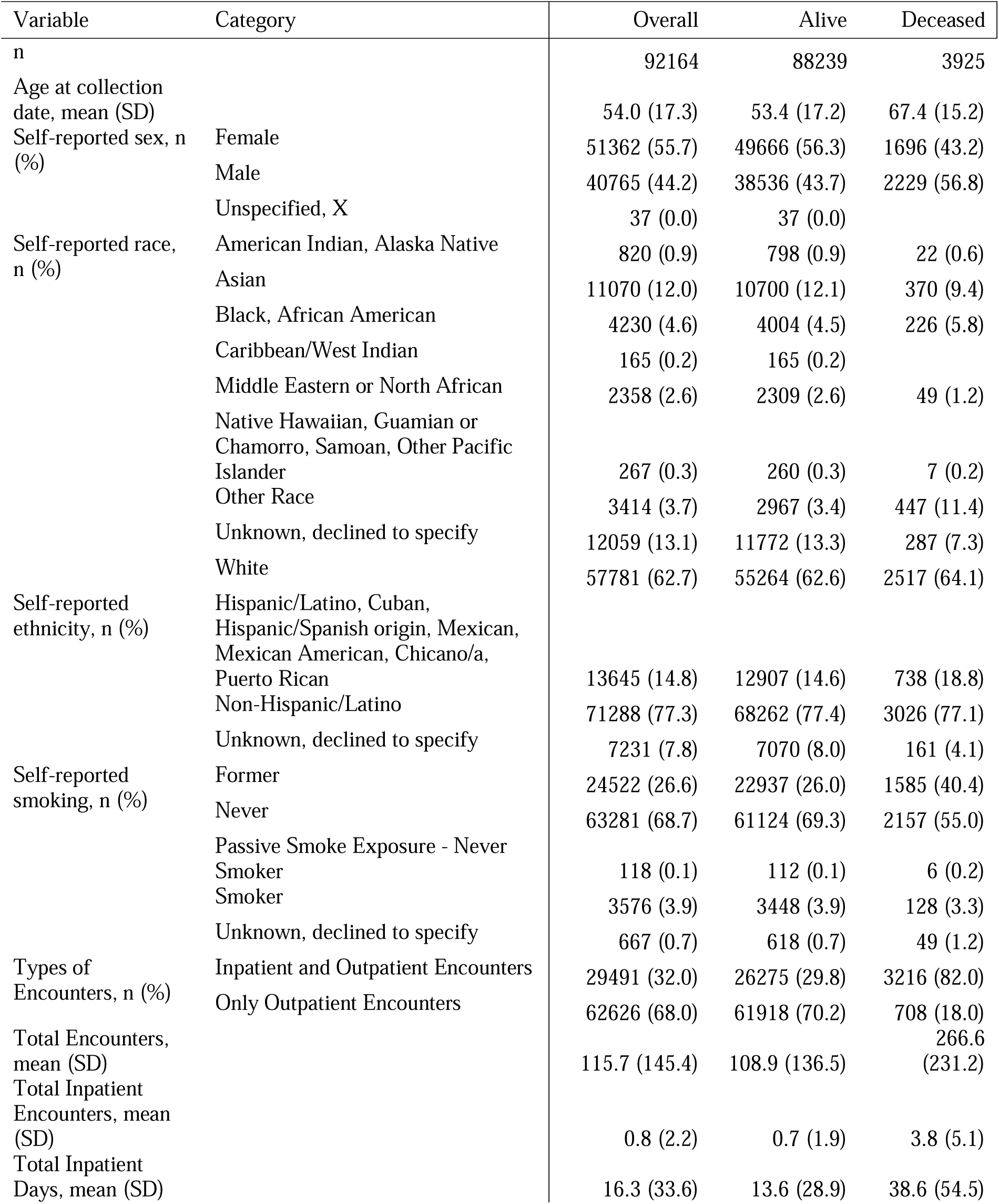
Baseline demographic information on the UCLA ATLAS population.

Although we primarily consider genetically determined ancestry in our analyses, rather than the social constructs of race and ethnicity^34,35^, we note substantial diversity based on participant self-reports. For instance, 62.7% of participants self-identify as White, 12% as Asian, 4.6% as Black or African American, 2.6% as Middle Eastern or North African, 0.9% as American Indian or Alaska Native and 0.3% as Pacific Islander. A substantial proportion (14.8%) report Hispanic or Latino ethnicity (**Table 1**).

### Disease burden varies by broad-scale genetic ancestry

Self-identified race/ancestry, a cultural-societal construct, and genetic ancestry are conceptually distinct^29,34,36^ and genetic ancestry must be considered to prevent confounding in genetic association studies^35,37^. We classified biobank participants into six broad-scale ancestry populations: European (EUR), African (AFR), South Asian (SAS), East Asian (EAS), Admixed American (AMR) and an Unclassifiable (UNC) population, aligning with the 1000 Genomes Project super-populations^38^ (**Figure 1e-g**; **Methods**). ATLAS is diverse relative to most other large biobanks (**Supplementary Note**). Consistent with self-reported race/ethnicity, almost a third of the biobank participants (32%) were assigned to non-EUR genetic ancestries. There was significant agreement between self-reported race and broad-scale ancestry – 99% of self-identified White participants were assigned to EUR or AMR populations (**Figure S3a**). Of those with “unknown” self-reported race, 46% were assigned to AMR ancestry and 45% to EUR.

We next asked how health system usage or disease burden varied by broad-scale ancestry, observing that the mean number of encounters varied significantly across ancestries (P-value = 7.5×10^-138^, ANCOVA, adjusted for genetic sex, age and Barriers to Accessing Services [BAS]^39^), with the highest numbers of total encounters in participants from AMR ancestry (adjusted mean = 16.2), followed by AFR (15.8) and SAS (14.2), with the lowest values in EUR (12.2) and EAS (12.9) (**Figure 1h**). We also assessed health burden using the comorbidity index score^40,41^, which was higher in EAS (adjusted mean = 3.7) and AMR (3.6) compared to others (all other populations 2.6-2.9; **Figure 1i**; P-value = 2.6×10^-5^, ANCOVA, adjusted for genetic sex, age, BAS and Area Deprivation Index [ADI]^42^). Similar results for disease distribution across ancestries were obtained after applying inverse probability weighting (IPW) to adjust for participation bias relative to the broader UCLA Health population (**Figure S3b**; **Methods**).

Increased encounter numbers were only partially explained by elevated comorbidity index scores, indicated by modest correlations between the two (mean total encounters *vs.* Elixhauser Comorbidity Index: R = 0.32, P-value <2.2×10^-16^; mean hospital encounters *vs.* Elixhauser Comorbidity Index: R = 0.26, P-value <2.2×10^-16^; **Figure S3c-d**).

The population diversity of ATLAS enables the interrogation of the combined genetic and social/environmental effects on the risk of disease diagnoses. To illustrate this, we assessed variation in medical conditions across populations, replicating multiple known associations (**Supplementary Note**; **Table S2; Figure S3e**). Further, we identified previously unreported associations (**Figure 1j**), including a lower risk of epilepsy in EAS compared to EUR participants (odds ratio [OR] = 0.46 with 95% confidence interval [0.32, 0.65], P_Bonferroni_ = 2.3×10^-4^), not previously detected^43,44^. Similarly, we found significantly reduced risk for bipolar disorder in those with AMR ancestry, clarifying conflicting findings in previous smaller studies^45,46^ (OR = 0.47 [0.40, 0.56], P_Bonferroni_ = 1.5×10^-17^). We also show a significantly lower risk of sleep apnea in EAS participants relative to EUR (OR = 0.67 [0.59, 0.75], P_Bonferroni_ = 4.1×10^-10^), which has been controversial^47–50^, with a weaker, but significant effect after correcting for body mass index (BMI) (OR = 0.84 [0.74, 0.95], P-value = 4.6×10^-3^; **Table S2**). These associations remained significant after adjusting for socioeconomic status (SES), using both the BAS and ADI measures, and with IPW (**Table S2**).

### Disease prevalence varies across fine-scale clusters

Fine-scale ancestries^37,51–53^, many of which are understudied^14–16^, reflect recent geographic or demographic stratification and can reveal important contributions to health disparities and disease risk^14^. We quantified fine-scale ancestry using identity-by-descent (IBD)^53,54^ in a population nearly three times larger than any previously published^52,53^. This approach identifies genomic regions shared between individuals due to a common ancestor and defines fine-scale ancestries, which we refer to as “clusters” (**Methods**)^52^. We identified 36 fine-scale ancestry clusters with at least 30 participants (**Figure 2a-b**; **Figure S4a-c; Supplementary Note**), labeled with both a numeric identifier (*e.g.*, IBD-01) sorted based on the cluster sample size, and a corresponding cluster-name to facilitate interpretation (**Methods**). Because PCA captures ancient population structure and IBD segments reflect recent shared ancestry, full agreement between these approaches is not expected, as is observed for some of our IBD clusters, such as IBD-02 (**Figure 2a**). We replicated previously identified clusters and added unreported ones, such as a Native Hawaiian cluster (n_IBD-29_ = 64) and a Bantu cluster (n_IBD-35_ = 32). The largest clusters consisted of Northern Europeans (n_IBD-01_ = 33,675) and Southern Europeans (n_IBD-02_ = 14,841). The remaining clusters represent the heterogeneity of ancestral origins in Los Angeles, including Ashkenazi Jewish (n_IBD-03_ = 14,262), Filipino (n_IBD-09_ = 1,438), Iranian Jewish (n_IBD-11_ = 707), and Armenian (n_IBD-16_,_IBD-23_,_IBD-31_ = 560) populations. The diversity of ATLAS is reflected not only by a relatively large portion of non-EUR individuals, but also by substantial diversity within the EUR broad-scale population (**Supplementary Note**). Several EUR clusters are large relative to published datasets, including Southern European, Ashkenazi Jewish, Armenian and Iranian Jewish^3,53,55–57^ (**Table S3**). Similarly, for EAS, we identified multiple clusters, including Filipino, representing the largest published Filipino genotyped cohort of which we are aware^58^. Despite this strength, for some populations, sample sizes are limited, which leads to relatively small clusters.

**Figure 2.**
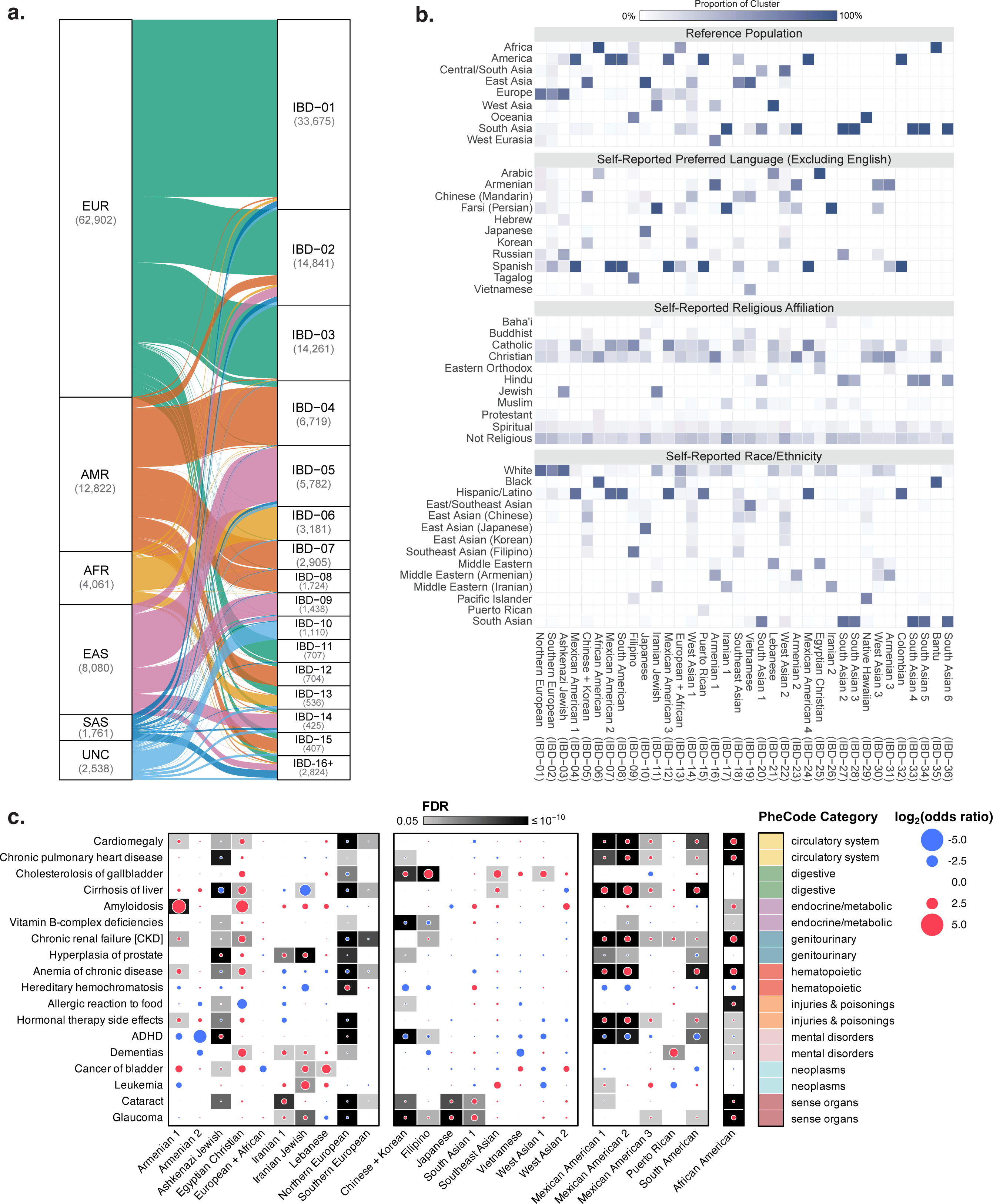
Identity-by-descent (IBD) mapping and fine-scale ancestry disease risk. **a.** Genetic ancestry assignments from broad- to fine-scale populations. **b.** Enrichment of reference populations, self-reported language, religion, and race across fine-scale ancestries. **c.** The risk of phenotype-defined phecodes across fine-scale genetic ancestries. Each fine-scale population was tested against all ATLAS participants outside of this population using Firth logistic regression across 1,253 phecodes.

For a nuanced understanding of diagnostic variation, we tested the prevalence of 1,253 phecodes^59–61^ across 24 fine-scale clusters with at least 100 participants. **Table S3** shows all associations and **Figure 2c** highlights selected associations (also available at https://atlas-phewas.mednet.ucla.edu/ancestry). We replicated known findings (**Supplementary Note**) and uncovered numerous associations between fine-scale clusters and diseases. For instance, Filipinos, an under-studied population in genetics research, exhibited the lowest risk for vitamin B-complex deficiency (OR_IBD-09_ = 0.50 [0.36, 0.67], FDR = 2.1×10^-4^), and the highest risk for cholesterolosis of the gallbladder (OR_IBD-09_ = 4.3 [2.9, 6.2], FDR = 5.6×10^-12^), both previously unreported. In Iranian Jewish subjects, another group with low research inclusion, we detected a previously unreported elevated risk for glaucoma (OR_IBD-11_ = 1.9 [1.5-2.4], FDR = 2.9×10^-6^), a condition known to be common in individuals of African and East Asian ancestry. We also observed several shared health risk patterns among Jewish ancestry clusters. Both Iranian and Ashkenazi Jewish clusters showed the highest risk for bladder cancer (Iranian Jewish: CI_IBD-11_ = 1.4-5.6, FDR = 0.02; Ashkenazi Jewish: OR_IBD-03_ = 1.5 [1.1, 1.9], FDR = 0.008) and a high hyperplasia of prostate risk (Iranian Jewish: OR_IBD-11_ = 2.3 [1.6, 1.8], FDR = 1.1×10^-13^; Ashkenazi Jewish: OR _IBD-03_ = 1.7 [1.6, 1.8], FDR = 4.6×10^-66^), and the lowest risk for cirrhosis of liver (Iranian Jewish: CI_IBD-11_ = 0.03-0.4, FDR = 3.1×10^-2^; Ashkenazi Jewish: OR_IBD-03_ = 0.4 [0.3, 0.5], FDR = 6.8×10^-24^). Mexicans and South Americans suffered consistently more from hormones’ adverse effects in therapeutic use, a previously unreported association (Mexican American clusters: OR_IBD-04,-07,and-12_ = 2.0-2.8 [1.3, 2.3], FDR = 1.6×10^-2^ - 2.1×10^-24^; combined South Americans: OR_IBD-08_ = 1.9 [1.4, 2.5], FDR = 1.3×10^-4^). These associations persisted after SES adjustment, using the BAS and ADI ranks. In most cases, associations held after applying IPW and also adjusting for SES, except for hormone adverse effects, where only the largest Mexican American group remained highly significant, likely due to smaller sample sizes resulting from the IPW analyses (**Table S3**). These findings demonstrate the utility of fine-scale ancestry in identifying underrepresented populations at risk for diseases.

We next focused on cardio-metabolic diseases due to their global impact on public health. We selected a few targeted cardio-metabolic phenotypes and compared disease risk across fine-scale clusters within the same broad-scale ancestry (**Methods**; **Figure S4d**). Strikingly, among Asian clusters, the Filipino cluster had an elevated risk for all tested cardio-metabolic conditions, which remained after adjusting for BMI. Across fine-scale EUR ancestries, the larger Armenian cluster showed high cardiometabolic disease risk. Iranian clusters, both Jewish and non-Jewish, had a relatively higher risk of coronary atherosclerosis, hyperlipidemia and type 2 diabetes, but not hypertension. Adjusting for SES factors and adding IPW maintained these significant patterns (**Table S3**), except abdominal aortic aneurysm in Filipinos, which lost significance, and hyperlipidemia in Iranian Jewish, which did not remain significant only when both SES adjustment and IPW were applied-likely due to reduced sample size, although the OR remained elevated.

### Ancestry-specific common variants associated with disease risk

To examine the robustness of ATLAS in predicting genetic risk and to assess the quality of our dataset, we first tested the utility of PGS in stratifying risk for common disorders. We observed high performance of PGS in EUR individuals – on average, 18.6% of patients diagnosed with major disorders were in the top PGS decile, a figure that rose to 41% for type 1 diabetes. Consistent with prior reports^62–65^, this predictive power was diminished in non-European ancestries (**Figure 3**; **Figure S5; Supplementary Note**).

**Figure 3.**
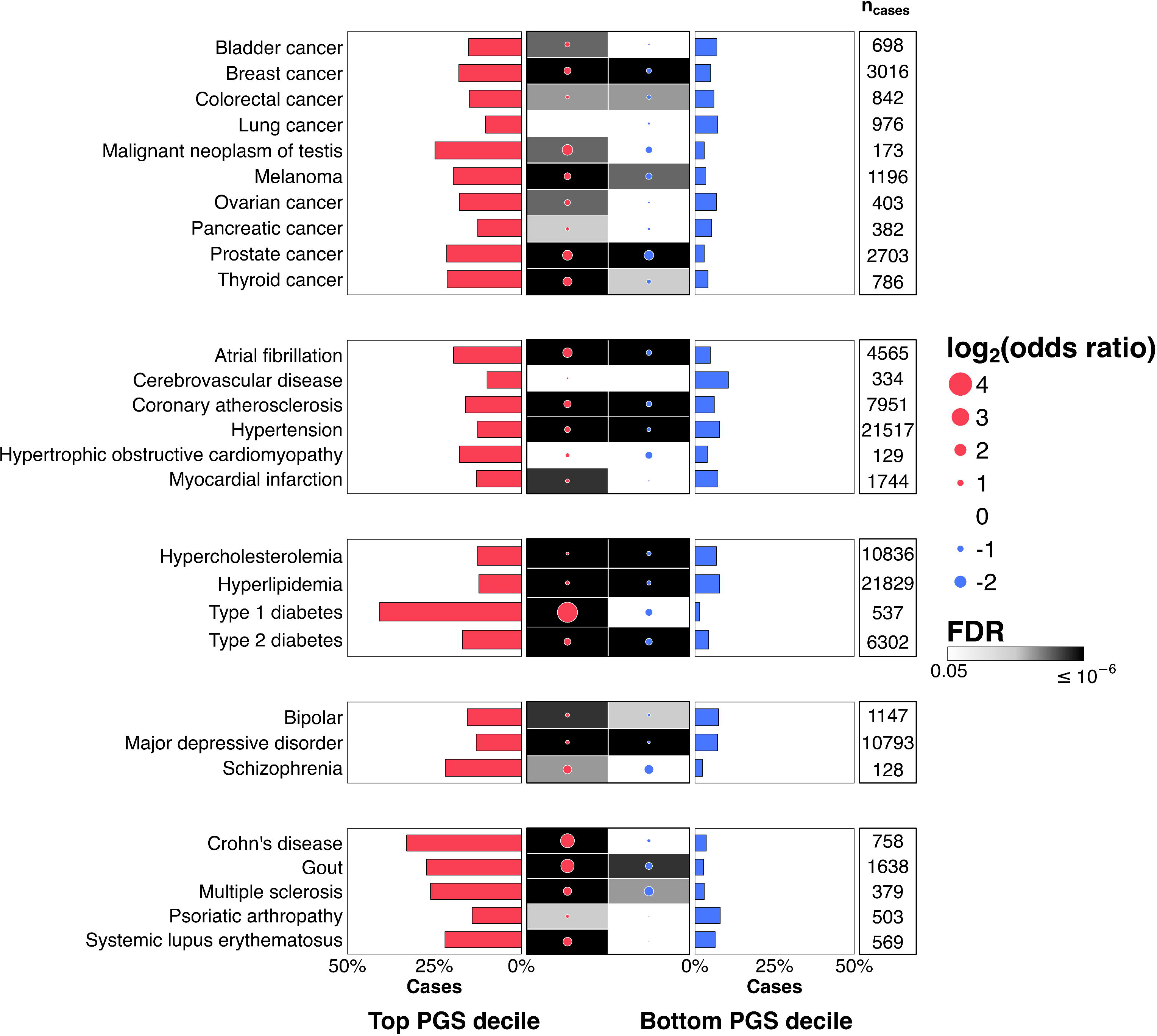
Polygenic risk and disease diagnoses. The top and bottom deciles of PGS were compared to the 5th decile to calculate the odds ratio (OR) using logistic regression (shown as dotmaps) in Europeans. The numbers of diagnosed patients assigned to the top or bottom PGS bins are shown in bar plots. Red represents the top PGS decile, and blue the bottom decile. Case counts are shown on the right. Diseases are divided into categories (top to bottom): cancer, cardiovascular, metabolic, neuro-psychiatric and immune/autoimmune. FDR was used for multiple testing correction.

Next, we performed phenome-wide association studies (PheWAS) using Regenie^66^ to map the common variant architecture of clinical diagnoses and laboratory measurements, conducting separate analyses within each of our broad- and fine-scale ancestral cohorts (**Methods**; n = 84,110 total participants). After linkage disequilibrium (LD) pruning, we identified 19,431 unique variant-phenotype associations passing a genome-wide significance threshold of 5×10^-8^ (**Table S4)**, with 5,772 passing the most conservative Bonferroni threshold of 6.4×10^-11^ (5×10^-8^ conditioned on 776 tested phenotypes). The majority (83.8%) of associations were detected in only one ancestry at genome-wide significance, though utilizing a more permissive threshold of 1×10^-4^ showed cross-ancestry support for a substantial portion of associations (54.7%; **Figure S6a-b**). Supporting the validity of the data used for PheWAS, we confirmed ancestry-specific differences in APOE allele frequencies and attenuated Alzheimer’s risk associated with the ε4 allele in African Americans and replicated known associations such as *PNPLA3* rs738409-G with non-alcoholic fatty liverin in AMR groups^67^ **(Supplementary Note**; **Figure S6c-e)**. We identified numerous ‘previously unreported findings’, which we defined as associations that have not been reported at genome-wide significance in the existing literature, the EMBL-EBI GWAS Catalog^68^, or summary statistics from UKBB^69^, Taiwan BioBank^7^, and AoU^5^. Notably, 39.5% of associations were unreported in the EMBL-EBI GWAS Catalog^68^ (**Methods**). We performed replication analyses for top previously unreported findings in independent biobanks, including AoU^5^, UKBB^69^, Taiwan BioBank^7^, and BioMe^8^ (**Table S4; Methods**). Summary statistics are available at atlas-phewas.mednet.ucla.edu.

We identified a previously unreported association between *FN3K* rs7208565-T and increased risk for intestinal disaccharidase deficiency across four EUR populations (**Figure 4a**; P_EUR_ = 3.80×10^-41^; OR_EUR_ = 1.26; MAF_EUR_ = 33.26%; MAF_IBD-01_ = 32.30%; MAF_IBD-02_ = 34.32%; MAF_IBD-03_ = 35.56%) and the AMR population (P_AMR_ = 4.90×10^-11^; OR_AMR_ = 1.31; MAF_AMR_ = 42.33%**; Table S4**). This variant was additionally associated with increased risk of the “other abnormal glucose” phecode (P_EUR_ = 1.73×10^-42^; OR_AMR_ = 1.25), while a variant in close LD (rs113373052-T) was associated with increased HbA1c (P_AMR_ = 1.40×10^-10^; β_AMR_ = 0.12), reflecting additional consequences on glucose homeostasis. Intestinal disaccharidase deficiency is the inability to completely digest sugars such as lactose and sucrose, leading to irritable bowel syndrome (IBS)-like symptoms^70^. *FN3K* phosphorylates glycated proteins, preventing the formation of advanced glycation end-products^71^. Supportive of these findings, rs7208565 has been previously associated with type 2 diabetes^72^.

**Figure 4.**
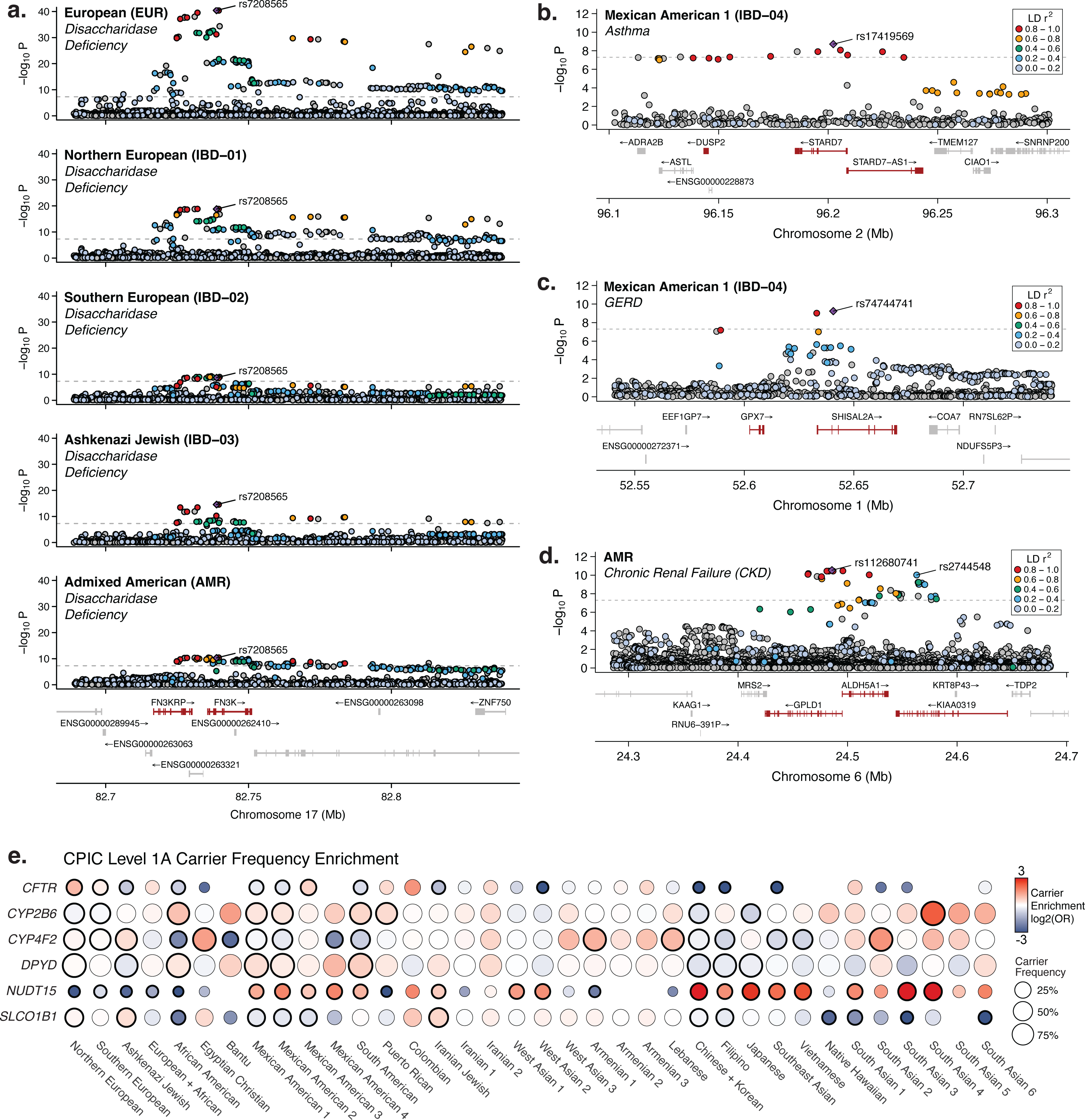
Selected PheWAS results and pharmacogenetic variant enrichment. a-d. Locus plots for unreported PheWAS associations between **a.** rs7208565-T and intestinal disaccharidase deficiency **b.** rs17419569-C and asthma. **c**. rs74744741-C and gastrointestinal reflux disease (GERD) and **d.** rs112680741-C/rs2744548-C and chronic renal failure. Point shading (blue to orange to red) indicates the level of ancestry-specific linkage disequilibrium (LD) with the lead single-nucleotide polymorphism (SNP), while gene shading (red) indicates prioritized risk genes. In **a-d**, Regenie’s Firth logistic regression was used for variant-trait associations. **e**. Pharmacogenetic variant enrichment across fine-scale ancestries. A bold border indicates FDR ≤ 0.05. Pharmacogenetic variants were aggregated for genes with multiple variants. Previously unreported enrichments highlighted in the text were for: *SLCO1B1* in Ashkenazi and Iranian Jews (OR_IBD-03_ = 1.4, [1.3, 1.5], FDR = 1.19×10^-35^; OR_IBD-11_ = 1.5, [1.2, 1.9], FDR = 5.00×10^-4^), *CYP4F2* in Ashkenazi Jewish (OR_IBD-03_ = 1.4, [1.3, 1.5], FDR = 1.36×10^-45^), Lebanese (OR_IBD-21_ = 2.0, [1.3, 3.1], FDR = 2.65×10^-3^), Armenian 1 (OR_IBD-16_ = 2.1, [1.6, 2.8], FDR = 9.07×10^-3^), Egyptian Christian (OR_IBD-25_ = 2.6, [1.4, 4.8], FDR = 2.97×10^-3^), and South Asian 2 (OR_IBD-27_ = 2.9, [1.5, 5.8], FDR = 1.56×10^-3^), and *PYD* in Mexican Americans, African American and South American (OR_IBD-08_ = 1.6, [1.4, 1.9], FDR = 3.41×10^-9^).

Furthermore, we discovered several previously unknown associations driven by low-frequency variants (MAF ∼1-2%) in non-EUR cohorts. For instance, we linked rs115750084-G in *DPP6*, a gene implicated in synaptic signaling^73^ and neurological disorders^74,75^, to an increased risk of major depressive disorder in the AFR cohort (P_AFR_ = 4.57×10^-8^; OR_AFR_ = 4.22; MAF_AFR_ = 1.09%; **Figure S6f**). In the EAS cohort, we associated rs77742325-G in *MAS1*, a key component of the bone-protective ACE2/Angiotensin-(1-7)/Mas axis^76^, with osteoporosis (P_EAS_ = 1.48×10^-8^; OR_EAS_ = 3.53; MAF_EAS_ = 1.79%; **Figure S6g**). We also linked an intronic deletion (rs202215133-ATATCATAG>A) in UBR5, an E3 ubiquitin ligase implicated in neuroinflammatory and neurodevelopmental syndromes^77,78^, with migraine headache in the Mexican American cluster (P_IBD-04_ = 1.66×10^-8^; OR_IBD-04_ = 5.58; MAF_IBD-04_ = 1.06%; **Figure S6h**). These associations were independently replicated, albeit with more modest effect sizes in the replication cohort (**Table S4**). Given that the variants are low MAF, further validation through direct sequencing in large cohorts will be essential to confirming their generalizability.

Similarly, our large AMR discovery cohort revealed several unreported associations. For instance, we linked *STARD7* rs17419569-C with increased risk for asthma in the Mexican American cluster (**Figure 4b**; P_IBD-04_ = 1.9×10^-9^; OR_IBD-04_ = 2.6; MAF_IBD-04_ = 2.5%). Prior work suggests that decreased *STARD7* expression is associated with enhanced allergic responses in the human lung and significant increases in airway hyperresponsiveness in haploinsufficient *Stard7* mice^79^, while rare variants within *STARD7* were linked with asthma (P_UKBB_ = 5.1×10^-3^; **Table S4).** In the same cluster, *GPX7/SHISAL2A* rs74744741-C was associated with gastrointestinal reflux disease (GERD) (**Figure 4c**; P_IBD-04_ = 5.8×10^-10^; OR_IBD-04_ = 1.6; MAF_IBD-04_ = 8.9%). *GPX7* has been associated with carcinogenesis in the context of GERD-associated Barrett’s esophagus^80^, while *SHISAL2A* has an unknown function, but is highly expressed in the small intestine and in lymphoid tissues^81^. Additionally, we identified two low-frequency AMR variants associated with chronic renal failure (CRF; **Figure 4d**), rs112680741-C (P_AMR_ = 2.94×10^-11^; OR_AMR_ = 3.78; MAF_AMR_ = 1.24%) and rs2744548-C (P_AMR_ = 9.59×10^-11^; OR_AMR_ = 3.17; MAF_AMR_ = 1.72%), nominating *GPLD1*, *ALDH5A1*, and *KIAA0319* as potential CRF risk genes. These associations did not replicate in available biobank populations (**Table S4**), possibly because the ATLAS AMR populations from the Los Angeles area represent distinct Mexican and South American ancestries^82,83^, whereas those from other biobanks (BioMe, for example) are comprised of individuals with distinct ancestral cluster assignments to Puerto Rico and the Dominican Republic. Other cohort-specific characteristics may also contribute to these differences, so independent replication in additional cohorts is needed.

### Frequencies of known clinically-relevant rare variants vary across ancestries

This first wave of the ATLAS WES catalog comprises more than 11.5 million autosomal variants, with a median of 9,821 missense and 159 loss-of-function (LOF) variants per individual, closely matching expectations from other studies^84^. Most (98%) WES variants were rare (MAF < 1% in any broad-scale ancestry), including 2,873,731 rare missense and 172,462 rare LOF variants, with a median of 15 rare LOF and 403 rare missense variants per participant (**Table S5**). AFR participants showed the greatest number of LOF and missense variants, matching prior results^85^. To evaluate our WES, we analyzed a predefined set of clinically relevant rare variants known to have elevated frequencies in specific populations and demonstrated that ATLAS captures these established patterns (**Supplementary Note**; **Figure S7a-e**). Then, we examined differences in 69 ClinPGx pharmacogenomic variants^86,87^ across fine-scale ancestries (**Figure 4e**). We identified several unreported enrichments in specific clusters, including a *SLCO1B1* variant, which affects the metabolism and toxicity of statins^88^ in Ashkenazi and Iranian Jews; a *CYP4F2* variant, rs2108622, that can increase the required dosage of warfarin^89^, enriched in Ashkenazi Jewish, Lebanese, Armenian 1, Egyptian Christian, and South Asian 2 individuals; and *PYD* variants, which affect the toxicity of chemotherapy drugs^90^, abundant in Mexican American, African American and South American clusters. This suggests the importance of considering fine-scale ancestry in precision drug prescribing.

Next, we used ATLAS’ ancestral heterogeneity to identify enrichments of known rare clinically relevant variants, aggregated by gene across fine-scale clusters to illustrate the utility of fine-scale ancestry in identifying populations at higher risk of rare, monogenic disease. We queried the full set of the curated rare pathogenic/likely-pathogenic (P/LP) ClinGen^91^ variants that have strong clinical and genetic evidence (n = 643), identifying 5,223 unrelated carriers (**Figure 5a**). Alongside known associations (**Supplementary Note**) within the Filipino cluster, we detected a previously unreported elevated carrier frequency for variants within the CDC Tier 1 gene, *LDLR*, that causes familial hypercholesterolemia (FH) (OR_IBD-09_ = 3.9 [1.4, 8.8], FDR = 0.03). This finding is notable given the high prevalence of dyslipidemia in Filipinos^92^ (**Figure S4d**). Indeed, LDLR FH variants accounted for 3.1 [0.3, 26.1]% (population attributable risk [PAR]) of high LDL cases (LDL > 190 mg/dl) in the Filipino cluster. However, due to the low number of carriers in this cluster (< 10), the PAR CI was wide, and independent replication is required to refine this finding. Additionally, we identified previously unreported elevated carrier frequency of variants that cause non-syndromic genetic deafness within two different genes in two separate clusters: (1) *MYO15A* in the Mexican American 1 cluster (OR_IBD-04_ = 6.9 [2.6, 17.4], FDR = 8.44×10^-4^) and (2) *CDH23* in the Chinese + Korean (OR_IBD-05_ = 6.0 [2.0, 15.6], FDR = 6.04×10^-3^) and Japanese (OR_IBD-10_ = 28.1 [8.9, 77.1], FDR = 7.38×10^-6^) clusters. We also identified previously unreported elevated carrier frequencies in the Ashkenazi Jewish cluster for Pendred syndrome variants in *SLC264A* (OR_IBD-03_ = 3.8 [2.9, 5.0], FDR = 8.61×10^-19^) and recombinase activating gene 2 deficiency in *RAG2* (OR_IBD-03_ = 2.9 [1.4, 5.8], FDR = 0.02). Finally, we focused on the clinically actionable American College of Medical Genetics and Genomics (ACMG)^93^ genes, where ClinGen-curated variants within these genes manifested a strong EUR bias (**Figure 5b**; **Figure S7f**; **Supplementary Note).** This bias was recently shown in AoU^23^ and we provide additional supporting evidence by demonstrating this bias in a single health system.

**Figure 5.**
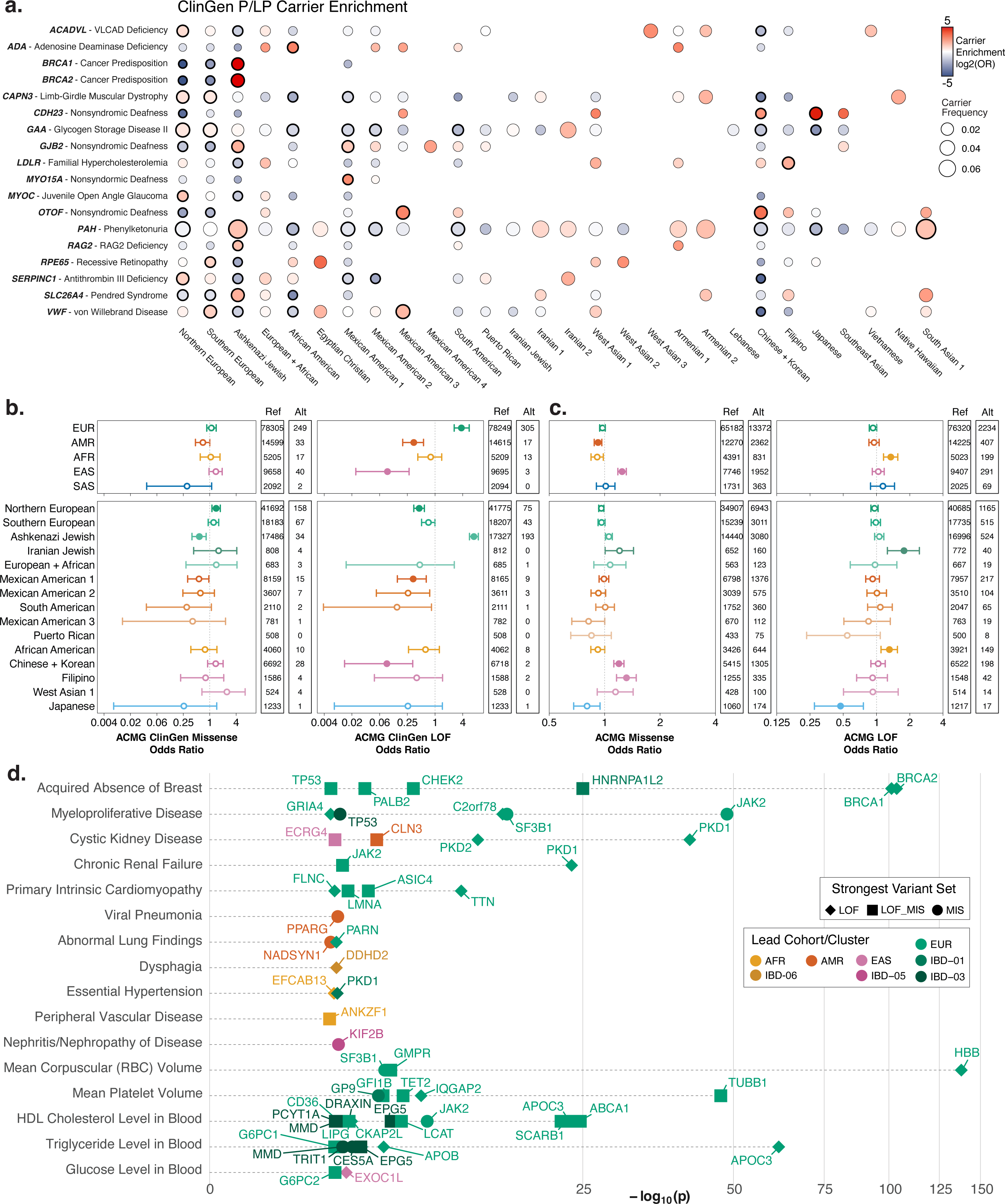
Rare variant findings. **a.** ClinGen pathogenic/likely-pathogenic (P/LP) variant enrichment across fine-scale ancestries. **b.** Differences across populations in the total numbers of rare ClinGene P/LP variants in American College of Medical Genetics (ACMG) genes. **c.** Differences across populations in the total numbers of rare predicted damaging missense and loss of function (LOF) variants in ACMG genes. In **b-c, ‘**Ref’ is the total number of reference alleles, and ‘Alt’ of alternative alleles, which are the rare P/LP ClinGen variants in **b**, and the rare, predicted LOF/damaging missense variants in **c**. In **a-c**, Fisher’s exact tests were used. In **b-c**, the top panels show broad-scale ancestries, and the bottom fine-scale ancestries. Filled points represent significant results (P_Bonferroni_ ≤ 0.05). **d**. Exome-wide association study significant results for selected traits/ancestries. Regenie’s omnibus "GENE_P" test was used for gene-trait associations.

### Predicted rare deleterious variants identify population-specific genetic risk

As clinical variant databases are primarily ascertained from EUR populations, we took a conservative approach to call rare predicted deleterious LOF (dLOF) and missense coding variants (dMIS). High-confidence dLOF were defined using the loss-of-function transcript effect estimator (LOFTEE)^94^, which accounts for biological context beyond protein truncation. dMIS variants were defined using a consensus of a majority (5 of 9) of state-of-the-art computational methods based on the Critical Assessment of Genome Interpretation (CAGI) project^95^ for pathogenicity prediction (**Methods**; **Figure S7g-h**). This approach is distinct from the one taken with ClinGen variants – variants identified in this manner may not have literature evidence for their impact on gene function but are potentially less biased by EUR oversampling.

First, we kept our focus on ACMG genes^93^, repeating the ClinGen-based analysis described above using predicted rare dLOF and dMIS variants. We examined the frequency of 1,602 rare dLOF and 6,487 rare dMIS variants within ACMG variants across populations (**Figure 5c**; **Figure S7i**). The results were consistent with previous work showing that AFR participants harbor substantially more dLOF compared to other ancestries^85^, and highlighted other groups with previously unreported low or high ACMG putative damaging variants for further investigation. Overall, these results contrasted in comparison with rare variants identified in ClinGen (**Figure 5b**), consistent with the interpretation that ClinGen variant frequencies are biased toward EUR participants, as is the case for most curated clinical datasets.

Second, to examine the influence of dLOF and dMIS on ATLAS phenotypes, we performed exome-wide association studies (ExWAS) across 17,537 protein-coding genes (**Methods**). Using Regenie’s unified gene burden association strategy^66^ we identified 1,099 unique Bonferroni-significant gene-trait associations. Within the EUR population, we replicated numerous known associations; 45 of the top 50 associations had been previously reported^84^ (**Table S6**). These include *PKD1* with cystic kidney disease and *TTN* with primary intrinsic cardiomyopathy (**Figure 5e)**^84^. We also identified numerous previously unreported associations (**Table S6)**. In the Northern European cluster, we detected gene-level associations between *HNRNPA1L2* and acquired absence of the breast (GENE_P_IBD-01_ = 1.0×10^-25^), breast cancer (GENE_P_IBD-01_ = 2.2×^10−8^), and malignant neoplasm of the female breast (GENE_P_IBD-01_ = 1.48×10^-7^). Notably, *HNRNPA1L2* is on chromosome 13, ∼20 megabases from *BRCA2*, representing a different locus. Though *HNRNPA1L2* has not been previously associated with breast cancer, it is highly expressed in breast invasive carcinoma^81^ and a paralog *HNRNPA1* was previously implicated in breast cancer progression^96^. In the Ashkenazi Jewish (IBD-03) cluster, we observed associations between *EPG5* with HDL cholesterol level (P_LOF_MIS_ = 3.6×10^-10^; β_LOF_MIS_ = 1.8 [1.2, 2.3]) and triglyceride level (P_LOF_MIS_ = 1.33×10^-8^; β_LOF_MIS_ = -1.81 [-2.4, - 1.2]). *EPG5* is an autophagy tethering factor classically associated with Vici syndrome^97^, a severe developmental disorder, though our findings suggest an additional role in lipophagy and lipid metabolism^98^.

In the AMR population, we identified an unreported association between dLOF and dMIS variation in *CLN3* and cystic kidney disease (GENE_P_AMR_ = 8.7×10^-9^), supported by experimental evidence that *CLN3* is highly expressed in medullary collecting duct principal cells and plays a role in osmoregulation^99^. We additionally identified unknown associations between *PPARG* and viral pneumonia (P_AMR,MIS_ = 1.1×10^-6^, OR_AMR,MIS_ = 53.0 [13.5, 208.5]), as well as *NADSYN1* and abnormal lung examination findings (P_AMR,MIS_ = 1.2×10^-5^, OR_AMR,MIS_ = 3.7 [2.1, 6.5]). *PPARG* is known to regulate macrophage response to pulmonary inflammation^100^, while *NADSYN1* knockout in mouse models was shown to cause abnormal lung development^101^, though neither has been previously associated with respiratory traits in humans.

Within the AFR population and African American cluster, we uncovered unreported associations between *DDHD2* and dysphagia (P_IBD-06,LOF_ = 1.2×10^-6^, OR_IBD-06,LOF_ = 8.3 [3.7, 18.7]) and *EFCAB13* and essential hypertension (P_AFR,MIS_ = 9.3×10^-7^, OR_AFR,MIS_ = 13.1 [4.5, 37.7]). These findings align with prior studies – variants in *DDHD2* have been shown to cause hereditary spastic paraplegia and symptoms of dysphagia^102^, though not in African Americans, while methylation studies have prioritized *EFCAB13* as a risk factor for heart failure^103^. We detected an additional association between *ANKZF1* and peripheral vascular disease (P_AFR,LOF_MIS_ = 1.6×10^-6^, OR_AFR,LOF_MIS_ = 58.8 [12.6, 275.1]), supported by recent experimental findings^104^.

Next, in the EAS population, we observed a previously unreported association between *AK7 a*nd non-rheumatic mitral valve disorders (P_EAS,LOF_ = 7.6×10^-7^, OR_EAS,LOF_ = 85.7 [39.2, 187.2]). While *AK7* has never been associated with cardiovascular phenotypes, it has a well-established role in primary ciliary dyskinesia^106^ and mutations in other ciliary genes have been associated with mitral valve prolapse^107^. We further identified an unreported association between *KIF2B* and nephritis/nephropathy in the Chinese/Korean cluster (P_IBD-05,MIS_ = 7.2×10^-7^, OR_IBD-05,MIS_ = 26.2 [9.0, 76.4]). Though *KIF2B* has not been linked to renal traits, other genes in the kinesin family (KIF) have been implicated in the pathogenesis of renal cancer^108,109^. All previously unreported findings described above were independently replicated in AoU^5^, UKBB^69^ or BioMe^8^ at a nominal P-value < 0.05 (**Table S6**).

Finally, we examined known risk associations to identify ancestry-specific patterns of deleterious rare variation. We tested two known *GBA1* rare missense variants, p.Glu365Lys and p.Thr408Met, in the AMR and EUR populations; both variants are reported to increase risk for Parkinson’s disease^110^. Despite similar allele frequencies within each population, p.Glu365Lys was only associated with increased risk in EUR (P_EUR,E365K_ = 0.01; P_AMR,E365K_ = 0.8), whereas p.Thr408Met was only associated with increased risk in AMR (P_EUR,T408M_ = 0.2; P_AMR,T408M_ = 0.003), displaying evidence of ancestry-specific risk stratification (**Figure S7j**). While we hypothesize that penetrance is modulated by population-specific genetic backgrounds, further studies are required to identify the causes of these differential associations. In total, we performed replication analyses for 21 top previously unreported PheWAS and ExWAS findings in independent biobanks and replicated 16 (76%) at a nominal P-value of 0.05.

### Longitudinal EHR data reveal genetic factors that shape GLP-1 receptor agonist efficacy

One of the advantages of EHR data is the ability to consider dynamic changes over time. To illustrate this, we integrated all main study components (genetic ancestry, common and rare genetic variants) to study the efficacy of GLP-1 receptor agonists (GLP-1 RAs) for weight loss. We focused on semaglutide, which had the greatest number of prescriptions in ATLAS (**Figure S8a-b;** 7,340 participants; **Figure 6a**). We observed a steady decrease in average weight up to ∼60 weeks of semaglutide treatment, consistent with previous findings^111–113^ (**Figure S8c**). We tested if the medication dose, route, sex, age, and initial BMI affect semaglutide efficacy, corroborating that dose and subcutaneous delivery versus oral delivery were positively correlated with weight loss (**Figure 6b**; linear mixed-effects model [LMM]: dose: P_Bonferroni_ = 3.3×10^-97^, semi-partial R² (explained variance) = 1.8%, effect size = -1.0; route oral *vs.* subcutaneous: P_Bonferroni_ = 7.2×10^-37^, semi-partial R² = 1.1%, effect size = 2.2). We did not detect a significant effect of age and sex on efficacy.

**Figure 6.**
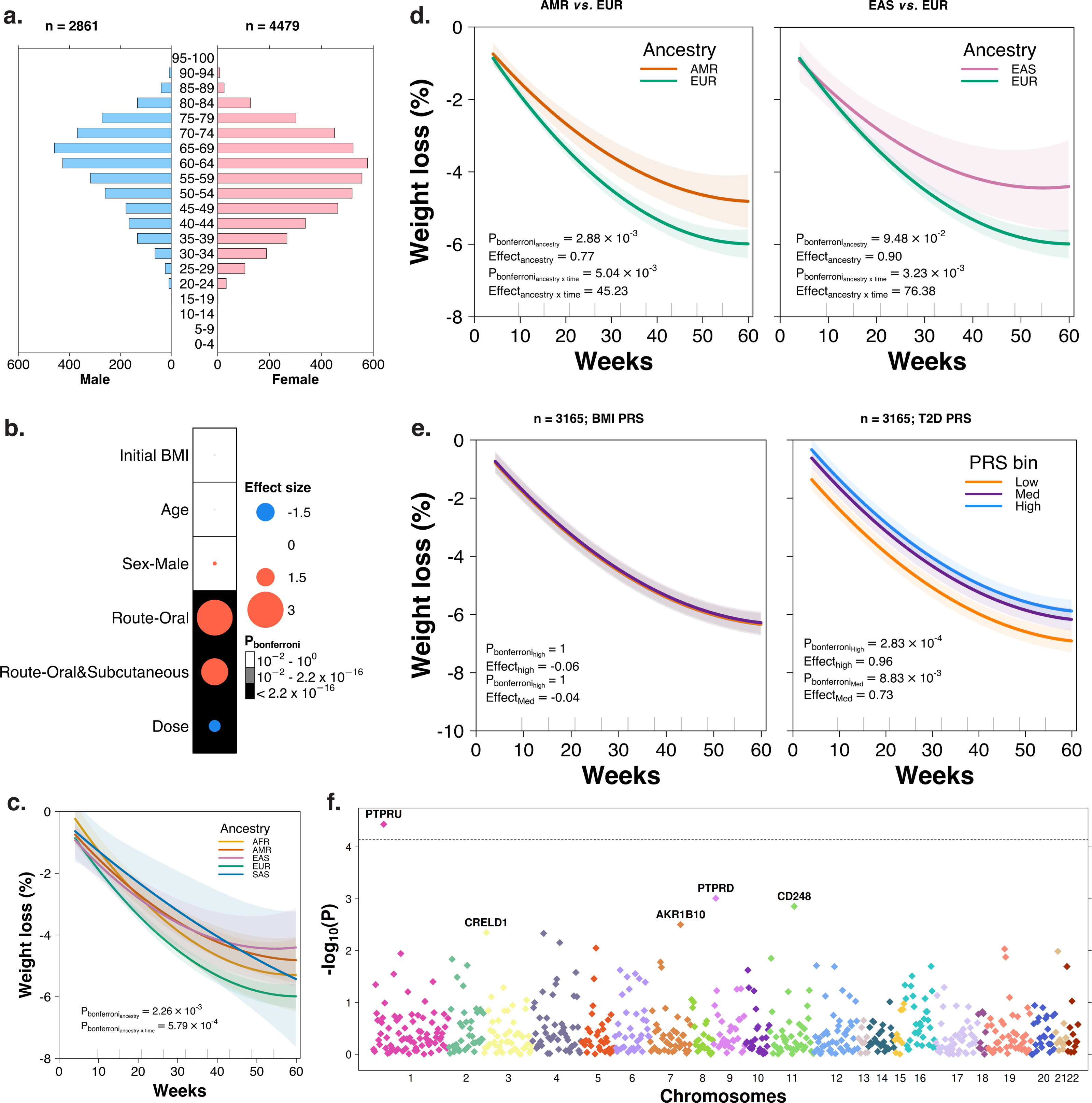
Semaglutide efficacy. **a.** Distribution of age by sex in semaglutide ATLAS users. **b.** The effect of baseline factors on weight loss in response to semaglutide. **c.** Weight loss patterns across genetic ancestry groups. **d**. Differences in weight loss patterns between AMR and EUR populations (left) and EAS and EUR (right). **e.** Relationships of body mass index (BMI) polygenic scores (PGS) (left) and type 2 diabetes PGS (right) with weight loss on semaglutide in EUR participants. Scaled PGS were divided into high, intermediate and low. In **b**,**d** and **e,** Bonferroni-adjusted P-values were obtained from a linear mixed-effects model that included covariates. In **c-e**, linear mixed model fitted values are plotted with 95% CI, based on longitudinal data with repeated weight measurements. **f.** Gene-level test results. Regenie additive model applied to test the genetic association between semaglutide-affected proteins and weight loss on semaglutide. The horizontal line marks the Bonferroni cutoff. The top 5 hits are labeled by gene names. For plotting, gene positions were defined based on the first base pair.

Next, we asked if semaglutide’s effects varied by broad-scale ancestry, finding that ancestry significantly influenced weight loss over 60 weeks of treatment (**Figure 6c**; ANOVA on a LMM; ancestry: P_Bonferroni_ = 0.002; ancestry×time: P_Bonferroni_ = 0.0006). Comparisons between EUR and the other populations revealed less weight loss in AMR, and a slower rate of weight loss in AMR and EAS (**Figure 6d**; LMM; ancestry_AMR_: P_Bonferroni_ = 2.9×10^-3^, β = 0.8; ancestry_AMR_×time: P_Bonferroni_ = 5.0×10^-3^, β = 45.2; ancestry_EAS_ ×time: P_Bonferroni_ = 3.2×10^-3^, β = 76.4), consistent with recent analysis based on a single time point, which showed reduced efficacy in AMR and AFR cohorts^114^.

Given limited evidence on the role of inherited factors in GLP1-RAs effectiveness^114^, we used ATLAS to explore the contribution of common genetic variation to semaglutide-related weight loss. We found no correlation between BMI PGS and semaglutide efficacy. However, weight loss was negatively correlated with DM2 PGS (**Figure 6e**; LMM, PGS_High_ *_vs._* _Low:_ P_Bonferroni_ = 2.8×10^-4^, β = 0.96; PGS_Med_ *_vs._* _Low:_ P_Bonferroni_ = 8.8×10^-3^, β = 0.73). The same relationship was observed in a simplified model using only maximum weight loss recorded^114^ (linear regression, P_Bonferroni_ = 1.3×10^-2^, β = 0.31; **Figure S8d**). We could not find an equally sized cohort for replication, but we used EUR AoU, which had a sample size 50% smaller than the ATLAS cohort used in this analysis with array genotyping (1,578 *vs*. 3,165 in AoU and ATLAS). In AoU EUR participants, we observed a concordant direction of effect, supporting replication of the finding, although statistical significance was not reached. This may reflect the smaller sample size or other population differences, such as in SES and compliance (**Figure S8e**). Larger samples are needed for more formal replication. As a second step, we conducted a genome-wide association (GWAS) meta-analysis across ancestries, but did not identify any significant loci (**Methods**; **Figure S8f**).

Finally, we used WES to test gene-level associations with semaglutide response (Regenie^66^; **Methods**). Given the modest sample size, we focused on EUR individuals and limited our test to the subset of proteins whose plasma abundance in humans was recently shown to be altered by semaglutide^115^. We identified a significant association of weight loss on semaglutide with *PTPRU* (P_Bonferroni_ = 7.6×10^-3^, β = -0.87) (**Figure 6f; Figure S8g**). For replication, we used AoU, with a cohort that was 21% smaller than the ATLAS cohort used (1,581 *vs*. 2,012 in AoU and ATLAS with WES). This analysis supported our original finding by showing a consistent direction of effect, although significance was not reached, possibly due to the smaller sample size or differences between the tested populations (AoU: β = -0.27, P-value = 2×10^-1^). Therefore, larger sample sizes in future studies will be needed to formally replicate this finding. However, the Bonferroni adjusted combined P from ATLAS and AoU remained significant, supporting this finding (P_Bonferroni_ = 2.3×10^−2^). This association involves 37 variants, of which one is common (rs2235937; nominally associated with weight loss; P = 9.7×10^-3^, β = -0.06 in EUR), and the rest are rare, the frequency of which varied across ancestries **(Figure S8h**; **Supplementary Note**). The direction of effect indicates that *PTPRU* activity is negatively associated with weight loss in patients taking semaglutide (variants in *PTPRU* with a predicted high or moderate functional consequence contribute to weight loss on semaglutide). Maretty *et al*. highlighted PTPRU as a protein whose abundance is elevated in individuals with higher genetic risk for increased BMI or type 2 diabetes and is downregulated by semaglutide treatment^115^. Although direct causality has not been established, these observations, together with our findings, support a model in which semaglutide-induced downregulation of *PTPRU* and genetically reduced PTPRU function promote greater weight loss during semaglutide treatment. This gene has no previous known functional relationship to weight loss or related metabolic functions. However, combined with the data from serum proteomics^115^, our genetic analysis nominates this protein kinase and its pathways as candidates for further investigation.

## Discussion

Large-scale, longitudinal population studies have accelerated our understanding of the causes and consequences of a wide variety of biomedical conditions. The Framingham study^116,117^ laid the foundation for population-based cohorts, followed by efforts like the UKBB and health system biobanks^3,8,9,11,53,72,84,118,119^. Despite the known sources of errors and incomplete phenotyping in health system EHRs, multiple studies have shown that many of these factors can be mitigated, allowing robust analyses^3,9,10,120^. Indeed, we were able to validate dozens of previously identified genetic associations in ATLAS based on EHR-derived phenotypes. We also leverage the heterogeneity of genetic ancestries in our population to validate and extend our knowledge of disease burden and genetic risk factors.

Prior work has mostly relied on utilizing broad-scale genetic ancestry to link ancestry to health risk^5,30,118^. Fine-scale ancestry discerns more recent genetic variation *via* shared IBD^121^. Through PheWAS and ExWAS across broad- and fine-scale ancestries, we identified numerous unreported genetic associations, many replicated. We leveraged fine scale ancestry definitions to benefit populations that are underrepresented in genetics and medical research^122^. For example, to our knowledge, ATLAS consists of the largest genetic dataset of Filipino individuals (n = 1,438 *vs*. 1,028 in the next largest dataset^58^), which can be used to improve precision care for this population and fuel disccovery. Similarly, our Ashkenazi Jewish cluster of 14,261 participants, which is 2.8 times larger than any prior single-study genetic cohort^57^, enabled previously unreported associations and enhanced risk quantification. Future work should assess the transferability of our results across populations^123^.

Discerning genetic variant risk frequency and effect, across fine-scale ancestries is powerful for optimized risk stratification and tailoring therapeutic intervention. It has been well demonstrated that diagnostic misclassification due to differences in variant frequencies across ancestries is a serious risk^22^. We emphasize that leveraging diversity within a single regional biobank can reduce confounding by ancestral genetic differences and non-genetic variables that differ across countries or geographically distinct biobanks. We demonstrated the application of longitudinal EHR to gain insights into health outcomes over time, by linking genetic factors to weight loss on semaglutide. This involved integrating dynamic EHR changes with detailed prescription information, including the medication dose and route, which we identified as major confounders. We considered only a single GLP-1RA drug, semaglutide, to avoid the complexity introduced by different GLP-1RAs, which have differing efficacy^124^. Lastly, our study highlights fine-scale populations with unreported low or high ACMG putative damaging variants for further investigation. We expect ATLAS to grow and become an engine for clinical intervention, allowing researchers to rapidly identify individuals for clinical studies and the implementation of precision medicine as the field continues to move forward.

### Limitations of the study

Certain limitations should be considered. First, due to the nature of EHRs, our study captures a partial view of patient phenotypes and care. Second, our analysis focuses on the risk of receiving a diagnosis for a disease, which is related, but not equivalent to the underlying risk of disease development. This may influence how our findings should be interpreted in the context of true disease incidence. Third, we relied on computational predictors for comparing abundances of predicted damaging variants in ACMG genes across ancestries. Some predictors may introduce ancestry-related biases^125,126^, potentially affecting the accuracy of the results. To mitigate these potential biases, we applied rigorous filtering and only retained variants upon concordant agreement in a majority of nine tools. Additionally, comparing the numbers of predicted damaging variants across ancestries may be less stable when small ancestry groups are involved. We therefore excluded groups with fewer than 400 individuals and reflected uncertainty using error bars. Lastly, while our study introduces multiple previously unreported associations, findings that could not be replicated should be further evaluated in independent cohorts as they become available.

## Data Availability

Due to privacy regulations/legal restrictions within the Health System and licensing terms, individual-level data cannot be published. Comprehensive processed data and summary statistics are available in the supplementary tables or through our browser (atlas-phewas.mednet.ucla.edu). Access to individual-level data may be possible through collaborative efforts and meta-analysis. This paper does not report original code.

## Resource Availability

### Lead contact

Questions should be directed to the lead contact, Daniel H. Geschwind.

### Materials availability

No materials were generated in this study.

## Acknowledgments

We are deeply grateful to the participants of the UCLA Health Biobank, to the DGSOM and UC Health for their support and development of ATLAS, and to Allen and Charlotte Ginsburg for their support of the UCLA Institute for Precision Health, which oversees ATLAS. WES data for this study were generated as part of the partnership of UCLA Health with Regeneron Genetics Center (RGC). R.H. was supported by EMBO Postdoctoral Fellowship ALTF 1131-2021 and the Prostate Cancer Foundation Young Investigator Award 22YOUN32. M.P.M. was supported by the UCLA-Caltech Medical Scientist Training Program (T32-GM008042) and by the National Institute of Mental Health (F30-MH135712). A.W. was supported by the National Human Genome Research Institute (F31-HG013462). J.F. was supported by the National Institute of Biomedical Imaging and Bioengineering Medical Imaging Informatics Training Grant (T32-EB016640). D.H.G., P.C.B., A.A.T.B. and C.L. were supported by UCLA CTSI. T.S.C. was supported by NIH grants K08AG065519-01A1, R01AG085518-01A1, U54NS123746 and California Department of Public Health, Chronic Disease Control Branch, Alzheimer’s Disease Program, under Contract #22-10079 and #24-10127. N.Zeltser. was supported by the National Human Genome Research Institute (T32-HG002536) and by the National Cancer Institute (F31-CA281168). V.A.A. and N.Zaitlen. are supported by the National Human Genome Research Insitute-R01HG011345.

## Author Contributions

D.H.G. obtained funding for the biobank and analyses; P.C.B., P.T.S., N.Zaitlen., B.P., A.A.T.B., V.A.A. and T.S.C. provided additional funding for analysis. D.H.G., P.C.B., P.T.S., and N.Zaitlen. designed, and E.E.K., B.P., A.A.T.B., V.A.A., and T.S.C. oversaw aspects of the study. R.H., J.F. and V.T. performed clinical data curation. T.N.Y., M.F.E.M., Y.P., N.Zeltser. and M.T. performed QC of the whole exome sequencing data. V.T., S.A., N.Zeltser. and R.H.-W. performed imputation of the array data, processing and QC. R.H., M.M., T.T., and V.P. performed curation and annotation of the WXS data. R.H. defined broad-scale ancestries, and A.W. fine-scale ancestries with support from C.C. and R.S. R.H., M.P.M., and J.F. performed the demographic investigation. R.H. performed analysis of PGS, clinical phenotypes and ancestries, ACMG variant differences, and semaglutide efficacy. M.P.M. performed PheWAS and ExWAS analyses, curated computationally predicted deleterious variants, and constructed the web portal. K.Q. performed locus pruning. L.O.C. assisted in setting up the web server. A.W. curated ClinGen P/LP variants and tested differences in frequencies of alleles across ancestries. J.A. provided statistical support. R.H., M.P.M., C.C. and S.L. performed replication analysis. R.H., M.P.M., A.W., P.T.S., and D.H.G. wrote and edited the first drafts of the paper, and all others edited subsequent versions. L.E., M.A., C.L., M.E.B., P.T. and D.B.M. were involved in building and maintaining the biobank, the data and its management. M.E.B., P.Thapaliya., P.Tung. and D.B.M. provided data curation, security, database and computer infrastructure.

## Methods

### EXPERIMENTAL MODEL AND SUBJECT DETAILS

The Institutional Review Board of the University of California Los Angeles gave ethical approval for this work. The IRB Number for the ATLAS Initiative protocol is IRB#17-001013.

Human participants included in this study were from the UCLA ATLAS Community Health Initiative. Enrollment procedures, consent, and data collection are described in Johnson et al^30^. Participants provided informed consent for research under IRB#17-001013. The cohort description is provided in the **Supplementary Note** and summarized in **Table 1** and **Figure 1**.

### METHOD DETAILS

Our study moves from characterizing the ATLAS cohort to ancestry-stratified analyses of disease burden – first at the broad-scale ancestry level, then at the fine-scale cluster level-followed by evaluations of common and rare genetic risk, and finally, an integrative case-study using longitudinal EHR data.

#### Array data and imputation

Array genotypes were obtained using the Global Screening Array. All data was mapped to GRCh38 and dbSNP, build 147^130^. Common haplotypes and variants were imputed using the TOPMed Freeze 5 panel^29,30^ using 668,127 observed single-nucleotide polymorphisms (SNP), resulting in a total of 50,757,223 high-quality called genotypes variants following imputation, an average of 2,048,050 per individual. Details regarding QC and the imputation procedure were described before^29^. Minimal QC was applied to the imputed genotypes. We retained only non-duplicated, bi-allelic variants with high imputation quality (R2 > 0.7), a minor allele frequency (MAF) > 0.1%, and a missing rate < 5%. Genotypes with a missing rate > 5% were excluded from further analysis. Concordance between genotypes determined by observed array variants and targeted Illumina sequencing was approximately 99.6%, determined using the vcf-compare command in VCFtools (v0.1.16)^131^.

#### Retrieving phenotype data

Demographic information, vitals, lab tests, International Classification of Diseases [ICD]) codes, and medication prescriptions were retrieved from the UCLA Data Discovery Repository (DDR), established on March 2, 2013^29^, containing deidentified participant EHR data from our health system.

Demographic, vital, and lab data were up to date as of Aug 24, 2024. For vitals and encounter counts, only records from in-person encounters (hospital visits, appointments, surgeries, office visits, and walk-ins) were considered. For vital signs, median values across all encounters were used to reduce the impact of potential recording errors or error-prone self-reported values. For lab data, the most recent values were considered. For **Figure 1c**, self-reported sex and age at the most recent time point at the time of analysis were used.

Yearly encounter numbers were retrieved in 2024. We included only complete yearly data up to 2023 to ensure consistency and avoid partial data from 2024. Hospital encounters during the COVID pandemic years (2019-2022) were excluded.

For associations involving clinical phenotypes, both ICD-9 and 10 were extracted. To ensure harmonized and comparable phenotypes across data sources, we adopted a structured, standard approach using phecodes^59–61^. We utilized the existing PheWAS catalog maps and applied standardized rules for case–control definitions. The ICD-9 codes were mapped to phecodes using phecode Map v1.2^59^, while ICD-10 codes were mapped using phecode Map v1.2b1^60^, both of which were obtained through the PheWAS catalog^61^. Cases were defined using the widely used “Rule of Two” approach^61,132–135^. For each phecode, participants were classified as cases if they had at least two occurrences of the same phecode, with occurrences spaced at least 30 days apart to ensure persistence of diagnosis. In total, there were 1,308 phecodes with at least 100 cases in ATLAS. Controls were defined as participants who had no record of the phecode in their EHR data and at least two recorded encounters in the system, following similar established rules for minimum data content^61,136,137^. Undecidable participants were excluded.

#### Phenotype validation

We used phecodes^59–61^ to define phenotypes and validated the generalizability of these phenotypes using several approaches. First, we tested cross-checked information between a variety of vital signs and lab tests and specific case-control groups defined by phecodes, using Wilcoxon tests and density plots, which confirmed the expected differences for these phenotypes. This included the following: 1) phecode-lab test pairs: Type 2 diabetes-Hemoglobin A1c, Hyperlipidemia-Trygllicoride, Vitamin D deficiency-Total 25-Hydroxy vitamin D, Deficiency anemias-Mean Corpuscular Volume (MCV), Deficiency anemias-Hemoglobin, and 2) the phecode-vital sign pairs: Essential hypertension-Blood pressure diastolic, Essential hypertension-Blood pressure systolic, Obesity-BMI, Anorexia nervosa-BMI, Short stature-Height **(Figure S2a-j**). Second, we replicated many known associations, including ancestry-related differences in disease prevalence for 28 major phenotypes. For example, across cancer types, we observed the highest risk for prostate cancer in AFR ancestry, stomach cancer in EAS, bladder cancer in AMR, and breast cancer in EUR^138^ (see more in the **Supplementary Note** and **Table S2**). Across fine-scale ancestries, we showed increased breast cancer and Crohn’s disease diagnoses in the Ashkenazi Jewish (IBD-03) cluster^53,139^ and observed the known increase in gout in the Filipino cluster (IBD-09)^140^ and Alzheimer’s disease and dementias in the Puerto Rican cluster (IBD-15)^141^ (see more in the **Supplementary Note**). We also replicated 11,756 variant–phenotype associations, such as a lower risk of Alzheimer’s disease for African American participants with the ε4/ε4 haplotype, and a decrease in MCV in response to *LUC7L* rs372755452-A in the broad-scale EAS ancestry. PGS prediction of 28 traits revealed significant enrichment in the top PGS decile for 89% of these traits in EUR (**Supplementary Note**). Collectively, the replication of known associations using ATLAS-defined phecodes indicates high-quality, well-defined phenotypes that match external definitions. Third, we used EHR data on cancer diagnoses to validate prostate and breast cancer cases (the two most diagnosed cancers) against phecode-defined case-control groups. We found a high agreement of 95%, for both cancers, considering decidable patients based on phecodes (**Figure S2k-l**).

#### ATLAS EHR baseline characteristics and their comparison to non-biobank UCLA patients

As all samples were collected incidentally, there was interest in characterizing the population, especially in comparison to non-biobank UCLA patients. To characterize the non-biobank UCLA patients while mitigating time-dependent confounding, we included a subpopulation with an encounter within one year of ATLAS launch date. Encounters could be either inpatient or outpatient, and we summarized the relative proportion of patients with only outpatient encounters or at least one inpatient encounter (there were no patients with only inpatient encounters).

To identify clinical phenotype patterns in ATLAS and to compare these to patterns of non-biobank UCLA patients, we used all ICD codes from each patient to encapsulate past and present conditions, subject to practical challenges^142^. For the ATLAS population, we captured a snapshot using their encounter closest to their biobank sample collection date. For the non-biobank patients, we used their closest encounter to the ATLAS launch date. These encounters represented the baseline encounters for both populations. The ICD codes at these encounters were converted to phecodes and phecode groups^59–61,143^ to represent meaningful categories of disease. Phecodes were extracted using pandas v2.2.2 with Python v3.9.19. We reported the unique phecodes with a prevalence ≥5% in the UCLA ATLAS population. ICD codes at the baseline encounters were also used to calculate Charlson and Elixhauser Comorbidity Indices ^40,41,144^, both measures that predict mortality. The scores were derived using the comorbidity R function v1.0.7^145^ with R v4.1.2. To compare the prevalence of disease categories between UCLA Biobank participants and non-biobank UCLA Health patients, we used logistic regression to test the association of each category with participant group, adjusting for age and sex.

In addition to characterizing patient populations at a baseline time, we also described differences in how they changed over time, an important consideration when assessing relative disease burden across populations. Participants were enrolled in UCLA ATLAS and were encountered within the health system at different times; therefore, we controlled the interval over which we measured change. We identified patients with at least one encounter between one and two years after their baseline encounter. From this subpopulation, we obtained patients’ first encounter within this window and summarized the cumulative encounters that occurred between the two times. We also used the ICD codes at the second encounter to derive new comorbidity scores, as well as extract updated phecodes. The change in comorbidity scores and increases in phecode prevalence provided insight into the evolving disposition of the UCLA ATLAS population over time.

#### Broad-scale genetic ancestry

The genetic ancestry of participants in the ATLAS dataset was estimated by assessing their proximity to the centroids of 1000 Genomes superpopulations in principal component (PC) space. PCA across common genetic variants demonstrates granular relationships and provides a quantitative basis for assessing relationships between ancestry and disease^29,36^. The top 20 PCs were calculated using the bed_projectPCA function (the bigsnpr^146^ R package v1.12.2) with default parameters. For every individual, the Euclidean distance to the centroids of the five broad-scale populations (AMR, AFR, EUR, EAS, SAS) was computed. Participants were assigned AMR or AFR ancestry if the nearest centroid corresponded to one of these populations, as these groups are well-separated in PC space. For EUR, EAS, and SAS ancestries, which exhibit more genetic overlap, a stricter distance threshold was enforced to minimize misclassification. Specifically, an individual was assigned to one of these ancestries if their distance to the nearest centroid was less than a scaled threshold, calculated as: Threshold=max(dist)×min(FST)/max(FST)×0.5, where max(dist) is the largest squared distance among centroids and min(FST)/max(FST) accounts for genetic differentiation. Participants who could not be assigned to any ancestry cluster under these criteria were labeled as "admixed/unknown." Visualization was performed using the Boutros Plotting General (BPG) R package v.7.1.0^147^.

#### Broad-scale ancestry related associations

Variation in encounter numbers across broad-scale ancestries was tested using ANCOVA, adjusted for genetic sex, age, and BAS rank to account for healthcare access differences. Comorbidity index values across broad-scale ancestries were compared using ANCOVA, adjusted for genetic sex, age, and both BAS and ADI ranks, which provide complementary measures of socioeconomic status. Since not all patients had BAS and ADI values, the sample sizes for these analyses were reduced (numbers are specified in the body of each figure). Adjusted means and CI were calculated using the R emmeans package, version 1.10.5^148^.

To test differences in disease diagnosis across ancestries, phecodes (retrieved as described in **Retrieving phenotype data**) were associated with genetic ancestry populations using logistic regression, adjusting for age (age at diagnosis for cases and the latest age for controls) and genetic sex if applicable. The following disease-phecode pairs were used to define disease diagnosis: Type 2 diabetes-Type 2 diabetes, Sleep apnea-Sleep apnea, Essential hypertension-Essential hypertension, Hyperlipidemia-Hyperlipidemia, Anxiety disorders-Anxiety disorders&Anxiety disorder&Generalized anxiety disorder, Asthma-Asthma, Parkinson’s disease-Parkinson’s disease, Type 1 diabetes-Type 1 diabetes, Schizophrenia-Schizophrenia, Crohn’s disease-Regional enteritis, Chronic kidney disease-Chronic kidney disease, Stage I or II, Multiple sclerosis-Multiple sclerosis, Major depressive disorder-Major depressive disorder, Cerebrovascular disease-Cerebrovascular disease, Atrial fibrillation-Atrial fibrillation, Hypercholesterolemia-Hypercholesterolemia, Hyperlipidemia-Hyperlipidemia, Coronary atherosclerosis-Coronary atherosclerosis, Hypertrophic obstructive cardiomyopathy-Hypertrophic obstructive cardiomyopathy, Myocardial infarction-Myocardial infarction, Systemic lupus erythematosus-Systemic lupus erythematosus, Gout-Gout, Bipolar-Bipolar, Psoriatic arthropathy-Psoriatic arthropathy, Epilepsy-Epilepsy, Neurofibromatosis-Neurofibromatosis, Dementias-Dementias, Obesity-Obesity, Obsessive-compulsive disorders-Obsessive-compulsive disorders, Autism-Autism, Migraine-Migraine, Alzheimer’s disease-Alzheimer’s disease, Coronary atherosclerosis-Coronary atherosclerosis, Posttraumatic stress disorder-Posttraumatic stress disorder. Patients under 18 years old, with ambiguous sex or with “unclassified” genetic ancestry were excluded. Visualization was performed using the BPG R package v.7.1.0^147^.

#### PGS analysis

PGS were calculated using array data after imputation in EUR participants. Related participants based on their genetic similarity were excluded (defined using PLINK v2.0a^109^ with the relatedness coefficient --king-cutoff 0.05). PGS were calculated using pgsc_calc^149^ with the default settings and --min_overlap of 0.65. Logistic regression was used to associate every PGS with the corresponding trait based on phecodes (see **Retrieving phenotype data**). The following PGS model IDs from the PGS catalog^150^ and their phecode pairs were tested: PGS002250-Malignant neoplasm of ovary, PGS003766-Cancer of prostate, PGS000004-Malignant neoplasm of female breast&Breast cancer, PGS001794-Thyroid cancer, PGS000079-"Melanomas of skin, PGS002264-Pancreatic cancer, PGS003395-Colorectal cancer&Colon cancer, PGS000729-Type 2 diabetes, PGS002025-Type 1 diabetes, PGS004254-Regional enteritis, PGS004699-Multiple sclerosis, PGS000134-Schizophrenia, PGS004760-Major depressive disorder, PGS001806-Malignant neoplasm of testis, PGS004687-Malignant neoplasm of bladder & Cancer of bladder, PGS000039-Cerebrovascular disease, PGS004526-Essential hypertension, PGS004706-Atrial fibrillation, PGS004784-Hypercholesterolemia, PGS002029-Hyperlipidemia, PGS003726-Coronary atherosclerosis, PGS000739-Hypertrophic obstructive cardiomyopathy, PGS004528-Myocardial infarction, PGS000803-Systemic lupus erythematosus, PGS001789-Gout, PGS002786-Bipolar, PGS000198-Psoriatic arthropathy. For prostate and testicular cancer, only males were included, and for breast and ovarian cancer, only females. An adjustment was made for age at diagnosis for cases and the latest age for controls, genetic sex when both sexes were included, and the first ten genetic PCs. For **Figure 3**, the top and bottom PGS deciles compared with the 5^th^ decile were considered, testing only EUR participants. FDR was used for multiple testing correction. The same process was repeated for other ancestries as presented in the supplementary material. Visualization was generated using the BPG R package v.7.1.0^147^.

#### Fine-scale ancestry pre-processing and quality control

##### Data

ATLAS array data were merged with genotyping data from the 1000 Genomes Project^38^, the Simons Genome Diversity Project^151^, and the Human Genome Diversity Project^152^. BCFtools^128^ annotate was used to harmonize variant reference SNP ID (RSIDs), and BCFtools^128^ norm with a GRCh38 genome reference was used to standardize the genotyping data. Sites or individuals with more than 1% missing were removed using PLINK^153^ --mind and - -geno. Only SNPs with MAFL>L1% across all participants were kept.

##### Phasing

SHAPEIT5^154^ with default parameters and the distributed GRCh38 map files were used to phase genotyping data, one chromosome at a time.

##### Identity-by-descent calling and processing

A custom Python script that converts PLINK bed files to PLINK ped/map^153^ files while conserving phasing information was used to convert genotyping data. Centimorgan data for the map files were generated using the same genetic map data in SHAPEIT5.

Identity-by-descent segments were called using iLASH^155^ with the following parameters: slice_size 350, step_size 350, perm_count 20, shingle_size 15, shingle_overlap 0, bucket_count 5, max_thread 20, match_threshold 0.99, interest_threshold 0.70, min_length 2.9, auto_slice 1, slice_length 2.9, cm_overlap 1 and minhash_threshold 55. Identity-by-descent was called one chromosome at a time.

##### Identity-by-descent quality control

Identity-by-descent segment outliers were removed as described in Belbin et al.^53^ and Caggiano et al.^52^. Segments overlapping centromeres or the human leukocyte antigen (HLA) region were removed. Regions that may have false positive identity by descent were identified using the following process and removed: total identity by descent per each SNP was identified by summing across all identity-by-descent segments that overlapped each SNP; SNPs with a total identity by descent greater than or less than three standard deviations from the genome-wide mean were removed.

##### Cluster identification

To identify clusters, we followed the approach of Caggiano et al.^52^ and Dai et al.^54^ and applied Louvain clustering^156^. An undirected network is generated based on pairs of individuals who share identity-by-descent segments: nodes are the individuals, edges are the total, genome-wide identity-by-descent shared as the edges. We used the Python package, NetworkIt^157^, to iteratively run Louvain clustering four times to detect fine-scale clusters.

##### Cluster merging

To avoid redundancy and maximize sample size, clusters were merged in two stages to produce a final set of consensus fine-scale clusters. First, following Caggiano et al.^52^and Dai et al.^54^, we computed pairwise Hudson’s F_ST_ using PLINK v2.0a^158^ across 378 clusters identified from the fourth layer of Louvain clustering. After removing clusters with fewer than 10 participants, to avoid unreliable F_ST_ estimates, we merged the remaining 356 clusters into 67 clusters if the pairwise F_ST_ was less than 0.001.

In the second stage, we refined clusters using IBD sharing. For each cluster pair, we examined all inter-cluster individual pairs to calculate: (1) IBD_mean_, the average cM shared, and (2) IBD_prop_, the proportion of pairs sharing at least 3cM of IBD as detected by iLash. We defined a composite metric for cluster merging, IBD_weighted_ = IBD_mean_×IBD_prop_, that captures both the extent and prevalence of genomic IBD sharing. Next, clusters were sorted from the smallest to the largest number of ATLAS participants. For each cluster, we computed the IBD_weighted_ score with all larger clusters and calculated the mean of these pairwise values. The cluster was then merged into the most similar larger cluster – *i.e.*, the one with the highest IBD_weighted_ score-only if that score exceeded the mean. Otherwise, the smaller cluster was retained independently. This process merged 14 small clusters into larger parent clusters, resulting in a total of 36 fine-scale clusters with ≥ 30 participants each for downstream analyses^52^. These fine-scale clusters were assigned unique identifiers (IBD-01 through IBD-36) and manually annotated with labels to ease interpretation.

##### Cluster labeling

We primarily relied on reference individuals, described in “Fine-scale Ancestry Pre-processing and Quality Control”, to add labels to clusters. When clusters did not contain reference individuals or were heterogeneous, we utilized patient-reported race, ethnicity, preferred language, and religious affiliation (in order of priority) to inform our cluster labeling. These aspects are not caused by identity-by-descent segment sharing but can be indicative of a shared culture for individuals within a cluster; these shared practices can influence a group’s demography, environment, and disease risk^159^. Our labels are not definitive and are our best attempt to generate informative assignments for each cluster.

#### Fine-scale ancestry and clinical phenotype associations

We used logistic regression to model the association between fine-scale ancestry assignment and phecode prevalence, estimating OR with 95% confidence intervals using the logistf R package v1.26.0 for Firth’s bias-reduced penalized-likelihood logistic regression^160^. Differences were tested between every fine-scale ancestry cluster with at least 100 participants (with no ambiguous genetic sex and over the age of 18) and all other ATLAS participants. For phecodes that are sex-specific, only participants of the corresponding sex were included in the analysis. Adjustment was made for age at diagnosis for cases, and the latest age for controls, and for sex when applicable. Phecodes with defined categories according to the PheWAS catalog^61^, and that were diagnosed in at least 100 participants with no ambiguous genetic sex and over the age of 18 in total in ATLAS were tested, resulting in 1,253 phecodes. In situations where the number of cases in a cluster was small, exact OR were not reported in the text to protect patient privacy. For visualization, fine-scale clusters were grouped into four panels (EUR, Asian, AMR, and AFR) based on the predominant broad-scale ancestry of participants. If the predominant ancestry was “Unclassified” (as in the Japanese and Egyptian Christian clusters), the second most prevalent broad-scale ancestry was used. The full phecode names presented in **Figure 2c** were: Hormones and synthetic substitutes causing adverse effects in therapeutic use, Cardiomegaly, Allergic reaction to food, Cirrhosis of liver without mention of alcohol, Vitamin B-complex deficiencies, Anemia of chronic disease, Cholesterolosis of gallbladder, Chronic renal failure [CKD], Dementias, Cancer of bladder, Cataract, Leukemia, Hereditary hemochromatosis, Attention deficit hyperactivity disorder, Glaucoma, Hyperplasia of prostate, Amyloidosis, Chronic pulmonary heart disease. These names were shortened in the figure and text for easier reading.

The same model was used to test differences in cardio-metabolic diseases using appropriate phecodes, for each fine-scale cluster within the same broad-scale continental ancestry. For this goal, clusters were assigned to broad-scale ancestries based on the predominant ancestry match among participants within each cluster. The largest population cluster within each broad-scale ancestry was used as the reference level for associations, adjusting for BMI, sex and age at diagnosis for cases, and the latest age for controls. As a sensitivity analysis, we tested whether the observed patterns persisted when correcting for SES factors (adjusting for ADI and BAS ranks) in addition to BMI, age, and sex. As a second step, we also added IPW (see **IPW analysis**).

FDR was used for multiple testing correction. The data were visualized using the BPG R package v.7.1.0^147^.

#### Comparison of ancestry sample sizes with published cohorts

The following studies were used to compare ATLAS’s broad-scale genetic diversity with other large scale biobanks (**Supplementary Note**): Halldorsson *et al*. (UKBB)^4^, Kurki *et al*. (FinnGen)^6^, Feng *et al*. (Taiwan Biobank)^7^, Zawistowski *et al.* (Michigan Genomics Initiative)^11^, Verma *et al*. (Geisinger MyCode)^161^, The All of Us Research Program Genomics Investigators *et al.*^5^, Shaw *et al*. (Vanderbilt’s BioVU)^162^, Verma *et al*. (Penn Medicine Biobank)^119^, Verma *et al*. (VA Million Veterans Program)^72^ and Wiley *et al*. (the Colorado biobank)^163^.

Studies that were used to compare sample sizes of ATLAS’s fine-scale ancestry clusters with other published cohorts with available genetic data (**Table S3**) included: Belbin *et al*. (BioMe fine-scale IBD clusters)^53^, Wu *et al*. and Li *et al*. (Ashkenazi Jewish)^57,164^, Haber *et al*. and Hovhannisyan *et al*. (Armenian)^55,56^, Mehrjoo *et al*. (Iranian)^165^, Larena *et al*. (Filipino)^58^, Sohail *et al*. and Ziyatdinov *et al*. (Mexican)^166,167^.

#### IPW analysis

IPW were calculated using logistic regression in R with ATLAS participants as cases and other UCLA Health patients as controls. As described above, we included UCLA Health patients who had at least one encounter within one year of the ATLAS launch date. The following covariates were included: self-reported race, sex, age group, BAS rank, and ADI rank. Individuals of unknown race were excluded. For each ATLAS participant, the probabilities were extracted from the model, and the weights were defined as 1 divided by the predicted probabilities. To assess whether previously unreported ancestry and disease status associations hold after applying IPW, we used the svyglm function (R survey package v4.4.8^168^) with a quasibinomial model, adjusting for age, sex, BAS rank, and ADI rank, including the calculated weights for ancestry groups with more than 5 cases. To compare comorbidity index values across broad-scale ancestries, considering IPW, linear regression was used, adjusting for age, sex, BAS rank, and ADI rank, including the calculated weights. Adjusted means and CI were obtained using the R emmeans package, v1.10.5^148^. Patients under 18 years old, with ambiguous sex or with “unclassified” genetic ancestry were excluded.

#### PheWAS

PheWAS were conducted using the Regenie v4.0 framework^66^ for all five broad-scale populations (AMR, AFR, EAS, EUR, SAS) and fifteen fine-scale clusters with at least 400 participants (IBD-01 through IBD-15). Related individuals were removed using a 0.05 kinship cutoff in PLINK v2.0a^158^. Sample sizes for each tested group can be found in **Table S4**. Association testing was performed separately within each group (*i.e.*, population or cluster) for 1,437 binary traits and 41 quantitative traits, including ICD-derived diagnoses and clinical laboratory measurements (see **Retrieving phenotype data**). Samples were restricted to those in predefined inclusion lists (--keep), and trait-specific covariates were provided *via* --covarFile, including age, sex (modeled categorically with –catCovarList sex), BMI, and the top 10 genetic PCs. Quantitative traits were divided into two analysis groups based on missingness patterns, following Regenie’s recommendation that traits with similar levels of missing data are modeled together in Step 1 (**Figure S12**).

##### Ridge regression (step 1)

For each group, unimputed array genotypes in bed format were filtered using PLINK v2.00a^158^ to produce a minimal set of variants suitable for estimating genome-wide polygenic effects through ridge regression in step 1 of Regenie. We included autosomal variants with call rate ≥99% (--geno 0.01), minor allele frequency ≥1% (--maf 0.01), and Hardy-Weinberg equilibrium p > 1×10⁻15 (--hwe 1e-15). Additional linkage disequilibrium (LD) pruning was applied using a sliding window of 1000 SNPs, advanced by 100 SNPs, with an r2 threshold of 0.9 (--indep-pairwise 1000 100 0.9), producing an average of 389,209 SNPs per group. For binary traits, step 1 was run with a minimum case count of 50 (--minCaseCount 50). For quantitative traits, step 1 was run with Rank Inverse Normal Transformation (--apply-rint) enabled to stabilize variance across lab values with differing distributions. For all traits, a block size of 1000 (--bsize 1000) was used, leave-one out cross validation (--loocv) was enabled, and sex was marked as a categorical covariate (--catCovarList sex).

##### Association testing (step 2)

Prediction files from step 1 for each trait and imputed genotypes in BGEN format were used to perform association testing in step 2 of Regenie. For binary traits, a minimum allele count of 20 (--minMAC 20) and a minimum case count of 50 (--minCaseCount 50) were required, and the Firth approximation was performed (--firth --approx) using a p-value threshold of 0.01 (--pThresh 0.01). For quantitative traits, a minimum allele count of 20 (--minMAC 20) was required and Rank Inverse Normal Transformation (--apply-rint) was applied. For all traits, a block size of 500 (--bsize 500) was used and sex was marked as a categorical covariate (--catCovarList sex). Locus pruning was performed separately for each broad- and fine-scale ancestry using Plink 2.0a to identify unique variant-phenotype associations.

##### Comparison to EMBL-EBI GWAS Catalog

To standardize phenotypes for comparison, phecodes for binary traits were mapped to the Experimental Factor Ontology (EFO) using the text2term v4.5.0 Python package. Each phecode was assigned up to three top-matching EFO terms. To assess the novelty of each unique variant-phenotype association, we queried the EMBL-EBI GWAS Catalog (June 27, 2025 freeze) for matching associations. An association from our study was classified as “previously unreported in the EMBL-EBI GWAS Catalog”, only if both of the following searches yielded no results. (1) Associations between the lead variant and any phenotype in the same phecode group as the associated phecode (*e.g.,* for colorectal cancer, we searched for associations with all EFO terms in the broader "neoplasm" category). (2) Associations between the nearest protein-coding gene (within 10kb) and any phenotype in the same category as the associated phecode.

#### WES sample preparation

Genomic DNA libraries were created by enzymatically shearing high molecular weight genomic DNA to a mean fragment size of 200 base pairs. Multiplexity of exome capture and sequencing was achieved by adding unique asymmetric 10-bp barcodes to the DNA fragments of single samples during library amplification. Equal molar amounts of DNA samples were pooled for exome capture using a slightly modified version probe library of xGen exome research panel from Integrated DNA Technology (IDT). After PCR amplification and quantification of the captured DNA, samples were multiplexed and loaded to Illumina sequencing machines for sequencing to generate 75-base-pair paired end reads. The samples in this study were sequenced using the Illumina sequencing machines, including NovaSeq 6000 with S2 or S4 flow cells and the NovaSeqX with 25B flow cells.

#### WES read alignment and variant detection

Sequencing reads in FASTQ format were generated from Illumina image data using the bcl2fastq program (v2.20, Illumina). Whole-exome reads alignment and germline small variant detection were conducted using the Original Quality Functionally Equivalent (OQFE) protocol described in Krasheninina et al., 2020^169^. Briefly, raw read files (FASTQ) were mapped to the GRCh38 reference obtained from https://ftp.1000genomes.ebi.ac.uk/vol1/ftp/technical/reference/GRCh38_reference_genome/ using BWA-MEM v0.7.17-r1188 in an alt-aware manner^170^. Duplicate reads were then marked with Picard v2.21.2^127^. The final CRAM files were compressed with SAMtools v1.2^128^. Germline variant detection was performed on each CRAM using a Parabricks accelerated version of DeepVariant v0.10.0 with a custom WES model^171^, resulting in a sample-level gVCF (genomic VCF). Per-sample gVCFs were merged with GLnexus v1.4.3^172^ into a joint-genotyped multi-sample project-level VCF (pVCF). Variant prediction was restricted to the exome capture region and the 100 base-pairs buffer on each side of the target regions. The WES was of high quality and reached an average coverage of 41.6-fold with a minimum of 24.2-fold, 90% having ≥ 34.7-fold in targeted regions, consistent with a previous publication^173^. The pVCF was converted to a PLINK file format using PLINK 1.9^153^ for downstream analyses.

These data underwent extensive quality control to ensure the absence of contamination, duplication, and other technical errors, as well as sufficient read depth to guarantee reliability and accuracy. Genetic duplicates were defined based on the aggregated genotype data of all sequenced samples. Sex was predicted based on the ratio of read coverage on chromosome Y over the whole exome read coverage. VerifyBamID v1.1.3^174^ was used to estimate sample contamination. Samples were excluded if they showed sex discordance, duplication, cross-indvidual contamination > 5%, or coverage < 20X in more than 20% off target regions. Overall, 386 samples were excluded for failing QC. Specifically, 249 samples were flagged for gender discordance, 12 had less than 80% of the exome covered at 20X, 80 showed contamination levels above 5%, and 74 were unresolved duplicates. After accounting for 29 samples that appeared in more than one exclusion category, the final number of unique samples excluded due to QC failure was 386. Alignment quality metrics were generated using Picard CollectHsMetrics v2.27.4^127^ and SAMtools stats v1.15.1 with default settings. MultiQC v1.27.1^175^ was used to systematically aggregate sample-level quality metrics. RTGtools v 3.12.1^129^ was used to assess variant QC metrics, including the number of variants for SNPs, small insertions and deletions; genotype counts; heterozygous-to-homozygous ratios for each variant type and transition/transversion (Ti/Tv) ratio. Genotypes were further confirmed using an alternative approach^176^ with 500 random samples, yielding a high concordance for on-target sites (median concordance of 99.5% for SNPs and indels).

#### WES summary statistics

For WES-based analyses, we used samples that also had array genotyping data (60,025 out of 61,797 patients with WES), which was necessary to consistently define broad- and fine-scale genetic ancestry. We annotated variants using Ensembl Variant Effect Predictor (VEP) v.112^177^ for 58,387 samples assigned to the EUR, AFR, SAS, EAS, or AMR ancestry class (*i.e.*, excluding unclassified genetic ancestry samples). Annotations were performed using the GRCh38 cache and the corresponding reference FASTA file. We limited our analysis to autosomal chromosomes to avoid technical artifacts in variant calling caused by the differences in ploidy between males and females, as well as the high-sequence similarity between the X and Y chromosomes in certain regions^178^. Counts for single-nucleotide variants, indels, multi-allelic, synonymous, missense, and LOF variants were restricted to whole-exome sequencing (WES)-targeted regions. Consistent with a previous UKBB WES study^179^, we classified LOF variants as those with the following consequences: stop_gained, start_lost, splice_donor, splice_acceptor, stop_lost, and frameshift. To increase reliability, we further restricted LOF variants to those flagged as high confidence by LOFTEE^94^. For multi-allelic variants, the predicted function for each alternate allele was determined using the --pick-allele option based on the default ordered set of criteria defined by VEP^177^. To determine the number of variants with MAF < 1% while accounting for ancestry-specific allele frequency differences, we retained variants with MAF < 1% in at least one ancestry group.

For multi-allelic variants, we defined the minor allele as the second most common allele (including the reference) and classified the variant as rare if the cumulative allele frequency of all alternate alleles was < 1%. For rare multi-allelic variants, we incremented a sample’s count only if it carried an alternate allele with MAF < 1%. In multi-allelic cases where the minor allele was the reference or the alternate alleles had AF ≥ 1%, we incremented a sample’s count if it carried any alternate allele. For rare functional variants (*e.g.*, missense), we incremented a sample’s count only if it carried an alternate allele with MAF < 1% corresponding to that functional category.

#### Replication of known elevated frequencies of rare P/LP variants

We identified variants *via* a literature search, where variants are known to be enriched within certain populations. For this goal we selected: 1) Familial Mediterranean Fever (FMF) related variants in the MEFV gene (V726A, M694V, M694I, M680I and E148Q)^180^ 2) the HBB:p.E7V variant, and 3) *PCSK9* P/LP variants^181^.

For FMF and *PCSK9*, carriers were identified as participants who carry at least one corresponding pathogenic variant within *PCSK9* or FMF variant group. For HBB:p.E7V, the frequency was estimated based on this variant alone. We only considered unrelated participants, using a kinship coefficient of 0.05 based on PLINK v2.0a^158^ king-cutoff. Fisher’s Exact test was used to test the carrier frequency of each cluster against the carrier frequency of the other clusters grouped together. The risk of amyloidosis (a common symptom of FMF) by carrier status was calculated considering carriers of known FMF and patients assigned the amyloidosis phecode (270.33).

#### Enrichment of ClinPGx and ClinGen P/LP variants

To test differences in frequencies of pharmacogenomic variants, 121 CPIC Level 1A variants were pulled from ClinPGx^86,87^. An overview of the analyzed variants can be found in the **Supplementary Note**. As variants spanned both common to rare frequencies, we used exome data for consistency. Overall, the variants were highly abundant in ATLAS, with 95.5% of all unrelated individuals carrying at least one. Of these variants, 69 were identified in the UCLA ATLAS exomes. To test enrichment of single alleles across clusters, “carriers” are identified as participants who carry at least one allele of a pathogenic variant within a gene associated with a monogenic condition (unless only a single variant was involved). We then calculated carrier frequency as the number of identified carriers over the total number of participants with WES available and were unrelated using a kinship coefficient of 0.05 based on PLINK v2.0a^158^ *king-cutoff*. To identify clusters with a significantly different carrier frequency of variants, we applied a Fisher’s Exact test to the carrier frequency of each cluster against the carrier frequency of the other clusters grouped together. We then used FDR corrections for p-values generated from all Fisher’s exact tests and selected significant clusters where at least 5 carriers were identified and FDR ≤ 0.05.

To identify clusters where participants were enriched for carriers of monogenic ClinGen variants associated with disease, we used pathogenic variants that were curated by experts within the field and underwent stringent review to be considered pathogenic from ClinGen^91^. ClinGen variants filtered to identify P/LP variants with autosomal dominant inheritance, autosomal recessive inheritance, or semidominant inheritance, resulting in 3,521 variants associated with 204 conditions overall according to ClinGen. We removed one *HNF4A* ClinGen variant due to a high allele frequency (MAF>0.01). Enrichment for variants, aggregated per gene, was tested as described above for ClinPGx variants with WES data. PAR was calculated for each cluster, showing significant enrichment of ClinGen P/LP variant frequencies aggregated by gene, focusing on genes associated with monogenic diseases with autosomal dominant or semidominant inheritance. To ensure sufficient power, we included only clusters with at least 30 individuals diagnosed with the disease with WES data available. We used the following formula: PAR = (p(RR-1))/(p(RR-1)+1), where p is the carrier frequency and RR is the ratio of the proportion of carriers with disease to the proportion of noncarriers without disease. For familial hypercholesterolemia, cases were defined as individuals with LDL levels exceeding 190 mg/dL, adjusted for statin use. In case of statin use, the LDL levels were divided by 0.7 if, as previously done^181,182^.

To calculate the PAR for breast cancer associated with the three Ashkenazi Jewish founder alleles, we used the formula above. Because individuals in this group are often aware of their genetic status and may pursue preventive breast cancer measures, we excluded individuals who underwent surgery (using the ‘acquired absence of breast’ phecode) but were not diagnosed with breast cancer, to improve the reliability of the estimate.

#### Differences in total ClinGen allele frequency within ACMG genes across ancestries

The list of all ClinGen^91^ P/LP variants identified in any ACMG secondary findings (SF) v3.2 genes^93^ was extracted as described above. Variants were labeled as missense or LOF, based on VEP v.112^177^ annotations. Missense variants were defined for ‘missense_variant’ variants according to the ‘consequence’ VEP output column. LOF was defined for high-confidence “HC” LOF variants based on LOFTEE^94^. All variants were rare across broad-scale ancestries. A WES plink file with all participants was filtered to include only the listed variants. Then, this file was broken into ancestry groups (*i.e.,* broad-scale population, or fine-scale cluster) using PLINK v2.0a^158^. Related participants were removed using a 0.05 kinship cutoff in PLINK v2.0a^158^. The frequency of each allele in every population group was calculated with the PLINK v2.0a --freq command, and total frequencies were summed up for rare missense and LOF variants separately. To test differences in the allele counts across populations, allele dosages were calculated with PLINK v2.0a^158^ --recode A option. Dosages were summed to calculate the total alternative (ALT) allele count within each population. The number of reference (REF) alleles was defined as twice the number of individuals in the group minus the number of ALT alleles. Fisher’s Exact tests were applied to test differences between the number of REF and ALT alleles in each population compared to all other participants not assigned to that specific group, considering only groups with over 400 participants. Bonferroni correction was applied for multiple testing correction. Plotting was done using the BPG R package v.7.1.0^147^.

#### Variant annotation using a consensus of computational tools

Variants from exome sequencing were annotated using VEP v.112^177^ with the dbNSFP v4.9a^183^ and LOFTEE^94^ plugins installed. The LOFTEE high-confidence “HC” flag was used to select for LOF variants predicted to have deleterious effects.

Missense variants were assigned a 9-point deleteriousness score based on a consensus of nine missense deleteriousness prediction toolkits, similar to methods described in prior biobank-scale rare variant studies^184^. We used the Critical Assessment of Genome Interpretation (CAGI) project^95^ to prioritize well-performing tools not trained on the same features. We selected five meta-predictors – ClinPred^185^, MetaRNN^186^, BayesDel_addAF^187^, VARITY_R^188^, REVEL^189^ – and four stand-alone predictors – AlphaMissense^190^, MutPred2^191^, VEST4^192^, ESM-1b^193^ – for use in our analysis. We assigned each variant a binary score per tool based on dbNSFP rank scores: 1 if the variant’s score exceeded the threshold score for being more likely a deleterious ClinGen or ClinVar variant than a background variant (**Figure S7g-h)**, and 0 otherwise. Summing these binary scores produced a deleteriousness score ranging from 0 to 9, with predicted damaging missense variants scoring ≥ 5 retained for downstream analysis.

#### ACMG putative damaging variant distribution across ancestries

A WES PLINK file was filtered to include computationally predicted LOF and predicted damaging missense variants (see above) in ACMG SF v3.2 genes^93^, providing a more comprehensive evaluation than the one based solely on ClinGen P/LP variants, which included only 17 genes. A WES plink file with all participants was filtered to include only the ACMG putative damaging variants, and the file was split into ancestry groups (broad- and fine-scale ancestries) using PLINK v2.0a^158^. Related participants were removed using a 0.05 kinship cutoff in PLINK v2.0a^158^. Allele frequencies were calculated with PLINK v2.0a, and only rare variants (MAF < 1% in all broad-scale ancestries) were kept. Allele dosages per individual were extracted using PLINK v2.0a, and the total rare LOF and predicted damaging missense alleles were counted separately per participant. First, the differences between the total numbers of REF and ALT alleles across ancestries were evaluated with a Fisher’s Exact test as described above (ClinGen ACMG analysis). Second, a Mann-Whitney U test with a Bonferroni correction was applied to test the difference in the distribution of rare LOF and predicted damaging missense counts per individual between any broad- or fine-scale group compared to all others, with groups that included at least 100 participants. **Figure S7e** presents the Mann-Whitney U test results, with statistically significant differences indicated by an asterisk (*). Visualization was done with BPG^147^.

#### ExWAS

ExWAS were conducted using the Regenie v4.0 framework^66^ for all five broad-scale populations (AMR, AFR, EAS, EUR, SAS) and fifteen fine-scale clusters with at least 400 participants (IBD-01 through IBD-15). Related individuals were removed using a 0.05 kinship cutoff in PLINK v2.0a^158^. Sample sizes for each tested group can be find in **Table S6**. Within each group, imputed genotype dosages in BGEN format (--bgen) and prediction scores from PheWAS step 1 (--pred) were used to conduct gene-based association testing for binary and quantitative traits across 17,676 genes. Samples were restricted to those in predefined inclusion lists (--keep), and trait-specific covariates were provided *via* --covarFile, including age, sex (modeled categorically with --catCovarList sex), BMI, and the top 10 genetic principal components.

##### Genotype QC (step 2)

Whole-exome genotypes in bed format were filtered using PLINK v2.0a, retaining variants with a call rate ≥ 90% (--geno 0.1), a minor allele count ≥ 1 (--mac 1), and Hardy-Weinberg equilibrium p-value > 1×10^-15^ (--hwe 1e-15). In addition, 784,548 variants overlapping low-complexity regions (LCRs) were excluded prior to analysis. Briefly, SNP positions were extracted from the .bim file, intersected using bedtools v2.29.1^194^ with annotated LCRs from the Genome in a Bottle Consortium^195^, and filtered from the genotype files, resulting in 12,326,160 exome variants retained for burden testing.

##### Gene-based association testing (Step 2)

Filtered whole-exome genotypes in BGEN format and prediction files from PheWAS step 1 were used to perform gene-based association testing. Regenie-style annotation, set, and mask files for predicted deleterious LOF and missense variants (see **Variant annotation using a consensus of computational tools**) were created programmatically using Python v3.11.9 with the polars v1.2.1 package. For binary traits (--bt), a minimum case count of 50 (--minCaseCount 50), a minimum minor allele count of 5 (--minMAC 5), and a block size of 1000 (--bsize 1000) were enforced. Firth logistic regression with saddlepoint approximation (--firth --approx) was applied for variants with p < 0.01 (--pThresh 0.01). For quantitative traits (--qt), rank inverse normal transformation (--apply-rint), a minimum minor allele count of 5 (--minMAC 5), and a block size of 500 (--bsize 500) were enforced. Regenie’s implementation of the RGC gene-based p-value test (–rgc-gene-p) was enabled for all analyses. Additionally, variants were binned by minor allele frequency using 1% bins (--aaf-bins 0.01), and SNP-level membership for each burden mask was recorded (--write-mask-snplist).

##### Variant-based association testing (step 2)

In addition to gene-based analyses, single variant association testing was performed for selected predicted deleterious LOF and missense variants using filtered whole-exome genotype dosages in BGEN format and prediction scores from PheWAS step 1. For both binary and quantitative traits, tests incorporated the same set of covariates (age, sex modeled categorically, BMI, and the top 10 genetic principal components) and enforced a minimum minor allele count of 5 (--minMAC 5). For binary traits, Firth logistic regression with saddlepoint approximation was applied for variants with pc<c0.01, along with a block size of 1000, while quantitative traits were evaluated using rank inverse normal transformed phenotypes with a block size of 500. A Bonferroni-adjusted p < 0.05 was defined as the cutoff for significance.

#### PheWAS and ExWAS replication in AoU, UKB, and Taiwan Biobank

##### PheWAS replication in AoU (logistic regression)

Replication analyses were conducted utilizing the Python Hail package v0.2.134^196^ on AoU Workbench using the Controlled Tier Dataset v8. Phenotypes were obtained by querying OMOP databases for per-patient ICD codes, which were subsequently converted to phecodes v1.2 utilizing the R PheWAS package v0.99.6^135^. Participants with both phecodes and single-read WGS data were filtered to remove participants flagged from genotype QC or from relatedness QC, resulting in 291,082 total participants. Logistic regression was performed within the given replication genetic ancestry cohort by testing case/control status for a given phenotype and SNP, utilizing age, sex, age^2, sex*age, sex*age^2, and genotype PCs 1-10 as covariates.

##### PheWAS replication in AoU (All by All)

Replications for variant-trait associations were obtained by querying the Controlled Tier Dataset v8 “All by All” allele count/allele frequency (ACAF) variant result tables using the Python Hail package v0.2.134 on AoU Workbench.

##### PheWAS replication in Taiwan Biobank

Replications for variant-trait associations were obtained by querying the summary statistics portal at https://taiwanview.twbiobank.org.tw/pheweb.php^7^.

##### ExWAS replication in AoU (All by All)

Replications for gene-trait associations were obtained by querying the Controlled Tier Dataset v8 “All by All” rare variant result tables using the Python Hail package v0.2.134^196^ on AoU Workbench.

##### ExWAS replication in UKB

Replications for gene-trait associations were obtained by querying the AstraZeneca summary statistics portal at http://azphewas.com/^69^.

#### PheWAS and ExWAS replication in BioMe

The BioMe Biobank consists of electronic health records and genetic data from approximately 60,000 participants from the Mount Sinai Health System in New York. Participant recruitment was between 2007 and 2023. This study was approved by the Icahn School of Medicine at Mount Sinai’s Institutional Review Board (Institutional Review Board 07–0529). All study participants provided written informed consent.

##### Genetic data

BioMe participants were genotyped using the Illumina Infinium Global Diversity Array (GDA; number of participants, N=23,430; number of variants, n=1,833,111) or Infinium Global Screening Array (GSA; N=32,595; n=635,623). Quality control consisted of removing participants with a call rate <95%, a mismatch between self-reported and genetic sex, and high heterozygosity. Duplicated sites and sites with a genotyping rate of <95% were removed. QC was done with PLINK2. Data was imputed with the TOPMED imputation server (Das et al) with genome build hg38. Genetic PCs were calculated across participants using PLINK2 for the genotyping sites after LD pruning. Exome sequencing data were generated by the Regeneron Genetic Center^197^. Quality control consisted of using the Goldilocks Filter^179^, and variants with quality scores <3 or depth of coverage scores <7 for SNPS, or <5 or depth of coverage <10 for indels were removed. Monomorphic sites were removed. Genetic ancestry was assigned using a random forest classifier trained on principal components derived from the 1000 Genomes data to align with the genetic ancestry assignment in AoU^5^. Participants were assigned a genetic ancestry using 10 PCs.

##### Phenotype data

Bio*Me* laboratory data were processed according to the QualityLab pipeline^198^. Briefly, quantitative lab values obtained in different clinical contexts (ambulatory, emergency, inpatient, and urgent care), were cleaned. Only labs with at least 100 patients and at least 1000 numeric observations were considered. Only adult (> 18 years) lab values were considered. Per lab, outliers were removed, defined as values as greater or less than 4 standard deviations from the mean of that lab. Non-numeric labs were removed. Labs must have >70% of the reported units matching. After cleaning, a median value and median age was calculated per person for each lab test in each clinical context and across all contexts. Laboratory phenotypes were manually assigned a match to UKBB phenotypes using available metadata.

Electronic health phenotype data in the form of ICD-10 codes were collapsed into PhecodeX phecodes^199^. Phecodes were transformed into a binary matrix, where each row was an individual and each column was a phecode. A participant had a 1 if they ever were diagnosed with that phecode, otherwise their value was set to 0. Age was calculated as current age, defined from 01/01/2025.

##### Association testing

Association testing was performed using Regenie v4.0^66^. Burden testing was performed using SKATO and the following masks: missense, missense_pLoF, pLoF, and synonymous. PLoF variants were defined using LOTFEE^94^.

#### Semaglutide investigation

##### EHR data curation

ATLAS GLP1-RAs prescriptions, including medication names, start and end dates, discrete dose, usage instruction, strength and route (oral or subcutaneous), were retrieved and grouped based on the following simple generic names: dulaglutide, semaglutide, liraglutide, exenatide, albiglutide and lixisenatide. For all statistical analyses, only semaglutide users were considered. Usage start dates were defined based on the earliest prescription start date for each patient. In the case of a missing start date, the prescription ordering date was used instead (in most cases, these two fields were identical). Overlapping medication periods were handled such that when a new prescription started, the previous one was considered to have ended. Similarly, missing end dates were determined using the start date of the next prescription when available. In the case of completely overlapping prescriptions with different instructions, the combination of both routes and the weighted average dose was considered. If the discrete dose information was missing, the medication dose was extracted from the instructions’ free text field. If the instructions also omitted the dose information, it was imputed for a given medication type, based on the weighted dose average from the entire cohort on the corresponding type. Initial weight and BMI were defined as the median of all available weight or BMI measurements recorded from in-person visits, within 6 months prior to the first prescription start date. Not relying solely on a single data point helps minimize the likelihood of typos and errors in the EHR. The percentage of weight change was obtained for every weight measurement recorded on in-person visits within the period of active prescriptions between 4-60 weeks in total on semaglutide. The medication dose at each time point was defined as the weighted sum of all medication doses (doses multiplied by the number of prescription weeks) by the measurement date. In case both oral and subcutaneous medications were used within a period, a combined route category was defined.

##### Statistical analysis

To identify the overall weight loss patterns across time, the FPCA R function from the fdapace package v.0.6.0^200,201^, which is suited to plot smoothed longitudinal data with repeated measurement, was used. This identified a consistent weight loss pattern up to 60 weeks, and sparse data points beyond ∼150 weeks. Thus, we restricted all analyses to this period.

For all association tests, only participants aged 18 years or older were included. For all analysis parts that involved longitudinal data with repeated measurements, a linear mixed model with the bobyqa optimizer and an increased function evaluation limit (maxfun = 10000) was used (lmer R function; the lmerTest R package v.3.1.3^202^). In each case, ANOVA was used to identify differences between models to define the best way to model a potential nonlinear relationship between weeks and weight loss, when treating weeks as a fixed effect, based on the Akaike Information Criterion (AIC). In some cases, the best model was achieved using a restricted cubic spline (RCS) with the rcs R function from the rms package (v.7.0.0), applied to weeks. In other cases, a polynomial function of weeks provided a better fit. To plot smoothed longitudinal data with 95% CI, the fitted model values (excluding covariates) and 95% lower and upper confidence bounds, were extracted using the visreg package^203^ v.2.7.0 and visualized using the BPG package^147^.

For testing the effect of baseline factors, fixed effects were defined for the medication dose, route, sex, age, initial BMI, first ten genetic PCs (excluding PC6 due to a strong collinearity with PC5 that disrupted the model convergence) and weeks on semaglutide. The model included both random intercepts and slopes for weeks on semaglutide. Bonferroni correction was applied to control for multiple testing.

For testing differences across ancestries, fixed effects were defined for the medication dose, route, sex, age, initial weight, a polynomial function of weeks on semaglutide, and an interaction between a polynomial function of weeks and genetic ancestry categorical class (EUR, AFR, EAS, SAS and AMR). The overall number of weight measurements was 24,145, with a mean of 5 repeated measurements per patient. Ancestry sample sizes were: European (EUR), 3189; African (AFR), 373; admixed American (AMR), 913; East Asian (EAS), 291; South Asian (SAS), 107. The model included both random intercepts and slopes for the polynomial function of weeks on semaglutide. An ANOVA was performed on the model with a Bonferroni correction to assess the global effects of ancestry and ancestry×weeks interaction. When a significant result was found, post hoc comparisons between groups were conducted using the summary lmerTest function (v.3.1.3^202^), applying a Bonferroni correction to all 12 class or class × time interactions.

To test the relationship between PGS and weight loss related participants based on their genetic similarity were excluded (defined using PLINK v2.0a^158^ with the --king-cutoff 0.05). Scaled BMI (PGS000027) and DM2 PGS (PGS000729) in EUR were divided into three equal bins each: low, intermediate and high scores. Then, using longitudinal data, for each trait, fixed effects were defined for the categorical PGS bins, medication dose, route, sex, age, initial weight, first ten genetic PCs (excluding PC6 due to a strong collinearity with PC5 that disrupted the model convergence) and a polynomial function of weeks. Random intercepts were defined to account for repeated measures within individuals. Bonferroni correction was applied to control for multiple testing. To test the relationship between PGS and weight loss, relying on a simplified model where the maximum weight loss was considered for each EUR participant, linear regression was applied. The model was adjusted for the medication dose and route, 10 genetic PCs, age, sex, and initial weight. Visualization was made with BPG^147^, with smoothed data and 95% CI using loess.as R function (fANCOVA package v.0.6.1^204^).

##### GWAS

The analysis was performed to test a relationship between the maximum weight loss on semaglutide and common genetic variants, for each broad-scale ancestry using SAIGE^205^. For step one, array observed variants were used following PLINK v2.0a^127^ filtering, with the flags: --maf 0.01 --mind 0.1 --geno 0.1 --hwe 1e-6. For the second step, array observed and imputed variants were used following plink filtering with --maf 0.01 --geno 0.05 --hwe 1e-6. A quantitative analysis was conducted with the traitType flag. The medication dose and route, the first five genetic PCs, age, sex, and initial weight were used as covariates. METAL^206^ was used for meta-analysis.

##### Gene-level tests

To identify genes genetically associated with weight loss, we used Regenie^66^ with an additive model for gene-level tests. Only EUR semaglutide users that are not related (relatedness coefficient --king-cutoff 0.05) with WES data were used (n = 2,012), and for each, the maximum weight loss record with the corresponding number of weeks was considered, using information on aggregate dose and route. The list of candidate genes was limited to Bonferroni-significant proteins whose plasma abundance was altered by semaglutide treatment^115^. We considered variants within these genes with a predicted moderate or high impact on the protein function, according to VEP v.1.2^177^. For step one, the variant list was limited to observed array SNPs following a PLINK v2.0a^158^ filtering with: --maf 0.01 --mac 100 --indep-pairwise 1000 100 0.9 --chr 1-22 --snps-only --geno 0.1 --hwe 1e-15. In step 2, we applied PLINK v2.0a filtering to variants across all EUR participants using the parameters --geno 0.05 and --hwe 1e-6. Subsequently, the file was filtered to include only semaglutide users and was used in Step 2. The medication dose and route, the first 10 genetic PCs, age, sex, and initial weight were used as covariates. Bonferroni correction was applied to control for multiple testing. Frequency of variants involved in *PTPRU* association with weight loss on semaglutide by ancestry can be found in the **Supplementary Note**.

##### Replication

For replication analyses in AoU the same processes and tools were conducted in EUR individuals. Controlled Tier Dataset v8 was accessed through the Researcher Workbench and imported in PLINK format, including exome variants in chromosome 1 (to extract *PTPRU* variants) and common ACAF variant datasets (for PGS calculation). Genetic ancestry and PCs derived from pre-defined assignments provided by AoU^5^. *PTPRU* VEP^177^ variant annotations were obtained using Python Hail v0.2.134^196^ to identify functional consequences. For the Regenie analysis, related individuals were not removed to maximise power, as Regenie is designed to account for relatedness. The combined P-value of ATLAS and AoU was calculated using Fisher’s combined probability test, implemented with the fisher function in the poolr R package v1.2.0^207^. Multiple testing was addressed using a Bonferroni correction that accounted for the number of gene-level tests in the discovery cohort, along with one replication P-value and one combined P-value.

### QUANTIFICATION AND STATISTICAL ANALYSIS

Statistical analyses were performed using R, PLINK, Regenie, and related tools. Logistic regression, linear regression, Firth-penalized regression, linear mixed models, and Fisher’s exact tests were applied in the relevant analyses, adjusting for appropriate covariates. Multiple-testing correction was applied using the FDR or Bonferroni adjustment, as specified in the relevant analyses. Detailed statistical models and software parameters are provided in the METHOD DETAILS section.

### ADDITIONAL RESOURCES

An interactive web portal enables users to explore the PheWAS results, examine fine-scale ancestry associations with clinical phenotypes, and download supplemental data at atlas-phewas.mednet.ucla.edu.

